# Discovery of novel obesity genes through cross-ancestry analysis

**DOI:** 10.1101/2024.10.13.24315422

**Authors:** Deepro Banerjee, Santhosh Girirajan

**Affiliations:** Bioinformatics and Genomics Graduate Program, The Huck Institute of the Life Sciences, University Park, PA 16802; Department of Biochemistry and Molecular Biology, Pennsylvania State University, University Park, PA 16802; Department of Anthropology, Pennsylvania State University, University Park, PA 16802

## Abstract

Gene discoveries in obesity have largely relied on homogeneous populations, limiting their generalizability across ancestries. We performed a gene-based rare variant association study of BMI on 839,110 individuals from six ancestries across two population-scale biobanks. A cross-ancestry meta-analysis identified 13 genes, including five novel ones: *YLPM1*, *RIF1*, *GIGYF1*, *SLC5A3*, and *GRM7*, that conferred about three-fold risk for severe obesity, were expressed in the brain and adipose tissue, and were linked to obesity traits such as body-fat percentage. While *YLPM1*, *MC4R,* and *SLTM* showed consistent effects, *GRM7* and *APBA1* showed significant ancestral heterogeneity. Polygenic risk additively increased obesity penetrance, and phenome-wide studies identified additional associations, including *YLPM1* with altered mental status. These genes also influenced cardiometabolic comorbidities, including *GIGYF1* and *SLTM* towards type 2 diabetes with or without BMI as a mediator, and altered levels of plasma proteins, such as LECT2 and NCAN, which in turn affected BMI. Our findings provide insights into the genetic basis of obesity and its related comorbidities across ancestries and ascertainments.

## MAIN

Obesity is a complex and heritable disorder that contributes to numerous comorbidities and poses significant public health challenges^1^. Obesity is influenced by a combination of genetic and lifestyle factors^2,3^, yet the genetic component underlying the etiology of obesity remains a key area of investigation. While large-scale studies have identified roles for common variants^4^, including significant effects of polygenic risk scores (PGS)^5^, and rare protein-truncating variants (PTVs) in obesity risk genes such as *MC4R* and *BSN*^6–9^, as well as protective genes such as *GPR75*^9^, these studies have predominantly focused on populations of European ancestry^6–8^. In fact, most recent obesity gene discoveries have been driven by population-scale rare variant analyses of the ‘white British’ population in the UK Biobank (UKB) cohort^10–12^, which represents a relatively homogeneous group of healthy aging adults from a specific demographic, shaped by an inherent survivor bias and a largely similar environment^13^. The consequences of this bias extend beyond gene discovery, affecting the generalizability of obesity-related findings to the broader global population and limiting the effectiveness of precision medicine approaches^14^. To overcome these limitations, heterogenous cohorts such as the All of Us (AoU) program, which more closely represents the general population, provide an opportunity to not only assess genetic effects in a different ascertainment context but also to identify novel associations^15^. *For example*, a recent multi-ancestry study identified rare PTVs in *UBR3* to be associated with multiple metabolic disorders by combining association statistics across biobanks, including UKB and AoU^16^. Such studies underscore the power of heterogeneous populations in expanding our understanding of the genetics of complex disorders.

Here, we conducted a rare variant association study of body-mass index (BMI) using genetic and phenotypic data from 839,110 adults from six continental ancestries, leveraging cohorts from the UKB and AoU initiatives. Unlike previous rare variant association studies of BMI, our study encompasses the widest range of ancestries to date, with the largest representation of non-European populations. By combining statistics across ancestries, we identified 13 genes, five of which have not been previously linked to BMI. While some genes showed consistent risk-conferring effects across ancestries, others showed European ancestry-specific bias, with significantly reduced effect sizes in non-European populations. These disparities highlight the critical need for more inclusive genomic studies for a complete understanding of the etiology and to ensure equitable interventions for obesity.

## RESULTS

### Cross-ancestry analysis to identify BMI-associated genes

We analyzed genetic and electronic health record data of 839,110 adults from the UKB and AoU cohorts representing six ancestries^15,17^ (**Table 1**). We used REGENIE v3.3^18^ to perform gene-burden association tests by collapsing rare (minor allele frequency <0.1%) protein truncating variants (PTVs, defined as predicted loss of function or deleterious missense variants) for each gene and measuring their effect on BMI independently for each ancestry across the two cohorts. The gene burden association studies for each ancestry across biobanks were statistically well-calibrated (λ_GC_=0.82 to 1.17) with genomic inflation rates comparable to previous rare-variant burden studies^6,12^ (**Supplementary Fig. 1**).

**Supplementary Fig. 1:**
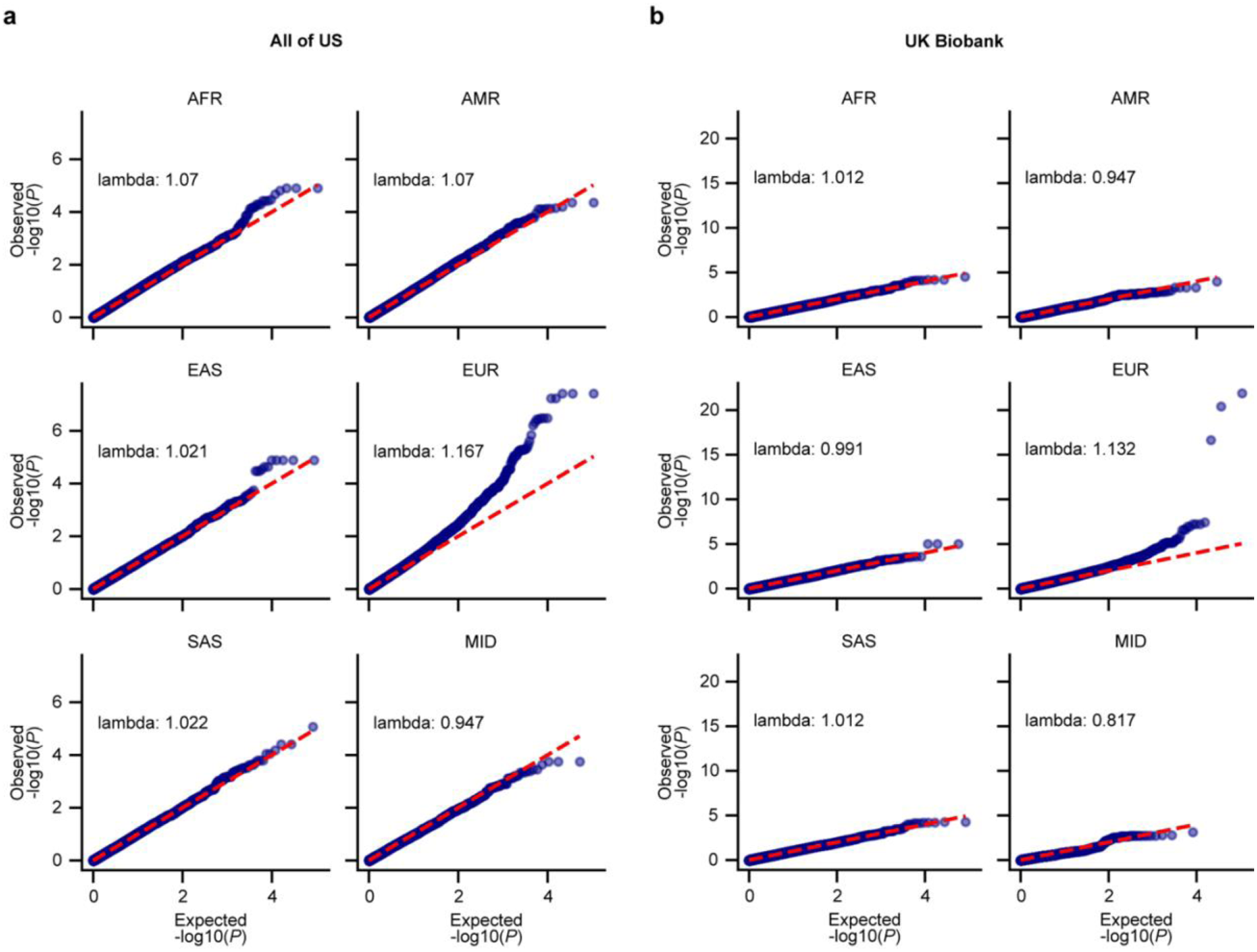
Genomic inflation rates of independent gene-burden tests. QQ-plot with genomic inflation rate (lambda) of ancestry specific gene burden association test for six continental ancestries in (**a**) AoU and (**b**) UKB. AFR: African, AMR: American, EAS: East Asian, EUR: European, SAS: South Asian, MID: Middle Eastern.

**Table 1:** Cohort and ancestry statistics analyzed in this study.

| Ancestry | UKB | AoU | Combined |
| --- | --- | --- | --- |
| African | 7,632 | 79,028 | 86,660 |
| American | 585 | 71,861 | 72,446 |
| East Asian | 2,297 | 9,390 | 11,687 |
| European | 435,335 | 217,581 | 652,916 |
| Middle Eastern | 90 | 1,435 | 1,525 |
| South Asian | 8,706 | 5,170 | 13,876 |
| **Total cohort (N)** | **454,645** | **384,465** | **839,110** |

Using individuals of European ancestry for discovery and those of non-European ancestry for replication, we performed an exome-wide inverse-variance weighted fixed-effect meta-analysis and identified 13 genes associated with BMI at exome-wide level of statistical significance (*P*<8.34×10^-7^, Bonferroni multiple testing correction for 20,000 genes and three variant collapsing models, see **Methods**) in both European and cross-ancestry meta-analysis (**Fig. 1, Supplementary Table 1**). Eight genes, including *MC4R*, *SLTM*, *APBA1*, *UBR3*, *BSN*, *PCSK1, UBR2,* and *BLTP1* have been associated with BMI or obesity in previous multi-ancestry or European ancestry-specific gene-based rare variant analysis^6,7,9,16,19^. We discovered novel associations for five genes with increased BMI, including *RIF1* (β=0.36, CI: 0.23, 0.50, *P=*9.05×10^-8^), *YLPM1* (β=0.36, CI: 0.24, 0.47, *P=*5.41×10^-10^), *GIGYF1* (β=0.29, CI: 0.19, 0.39, *P=*4.3×10^-9^), *SLC5A3* (β=0.15, CI: 0.10, 0.21, *P=*1.90×10^-7^), and *GRM7* (β=0.12, CI: 0.08, 0.17, *P=*2.25×10^-7^) with effect sizes comparable to known obesity genes (**Fig. 1**). Re-analyses after adjusting for genomic inflation using METAL^20^ retained significant associations for all genes, except *SLC5A3* (*P*=1×10^-6^) and *GRM7* (*P*=1×10^-6^) that only reached suggestive significance (**Supplementary Table 2**). The rare variant burden associations for all five novel genes were independent of any nearby common-variant BMI associations (at *P*<0.01), with effect sizes consistent with our original analysis (**Supplementary Table 3, see Methods**). We also performed leave-one-variant-out sensitivity analysis and found that the effect on BMI is not driven by a single PTV, but rather by any of the PTVs in these genes (**Supplementary Table 4**).

**Fig. 1:**
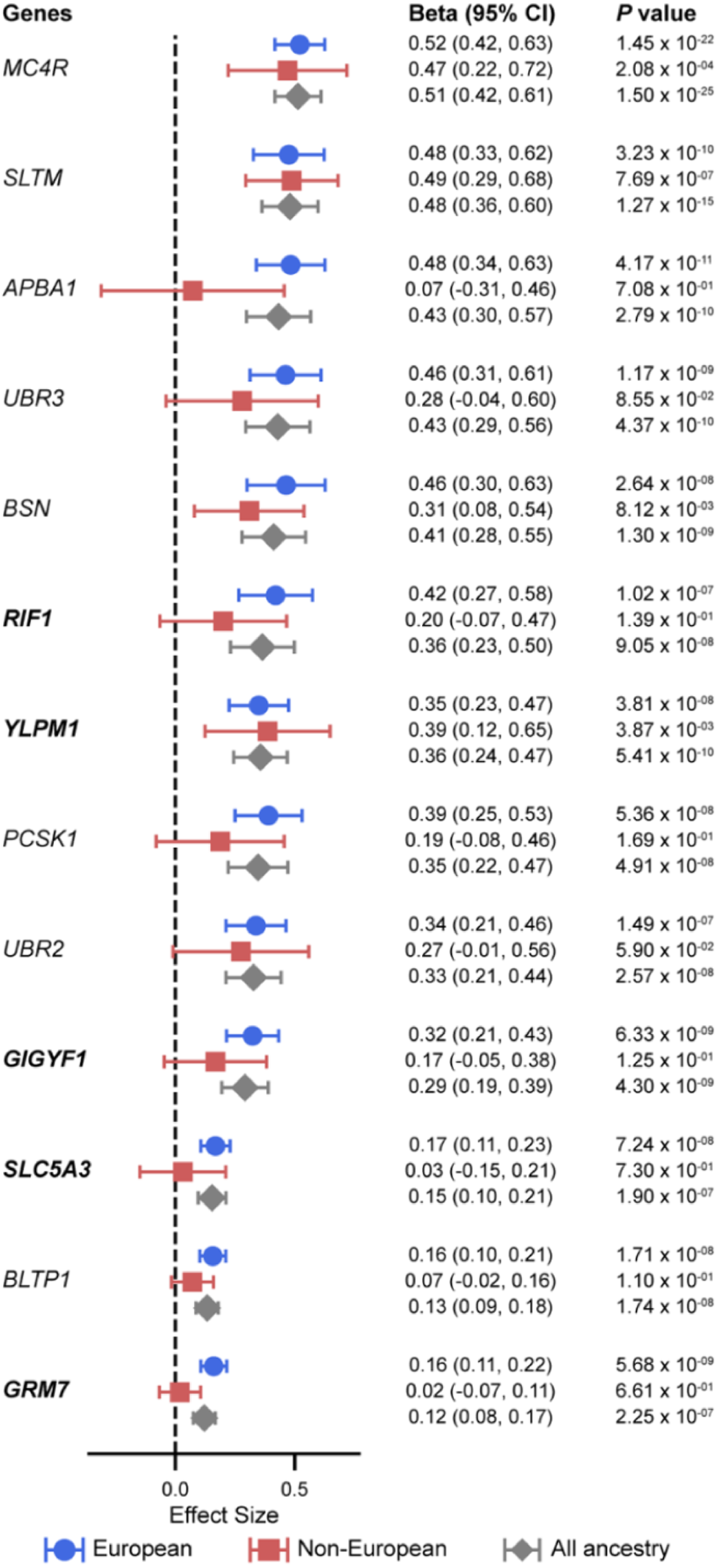
Meta analysis results of BMI associated genes. Effect sizes along with 95% confidence intervals (CI) and significance values (*P* value) in European, non-European, and combined meta-analysis for BMI associated genes. Three variant collapsing models were tested for each gene and the most deleterious mask that reached exome wide significance is shown here. Novel BMI associated genes are indicated in bold letters. Data for all genes contributing to changes in BMI are provided in **Supplementary Table 1**.

The effect of *YLPM1* on BMI was consistent in both European (β=0.35, CI: 0.23, 0.47, *P=*3.81×10^-8^) and non-European (β=0.39, CI: 0.12, 0.65, *P=*3.87 x 10^-3^) subgroups, with no evidence of ancestral heterogeneity (**Fig. 2, Supplementary Tables 1and 5**, Cochran’s Q=0.06, *P*=0.80). Such consistency in effect sizes between the two subgroups was observed only for *MC4R* (Cochran’s Q=0.14, *P*=0.71) and *SLTM* (Cochran’s Q=0.01, *P*=0.92) among the known obesity genes. On the other hand, *RIF1*, *GIGYF1*, *SLC5A3*, and *GRM7* showed about two-fold reduction in effect sizes in the non-European group, with *GRM7* also showing evidence of significant heterogeneity (**Supplementary Fig. 2, Supplementary Tables 1and 5**, Cochran’s Q=7.33, *P*=0.006). We also observed similar reduction in effect sizes in the non-European group for known obesity genes, such as *APBA1*, *PCSK1*, and *BLTP1* as well, with *APBA1* showing significant heterogeneity between European and non-European groups (**Supplementary Tables 1and 5**, Cochran’s Q=3.85, *P*=0.05).

**Fig. 2:**
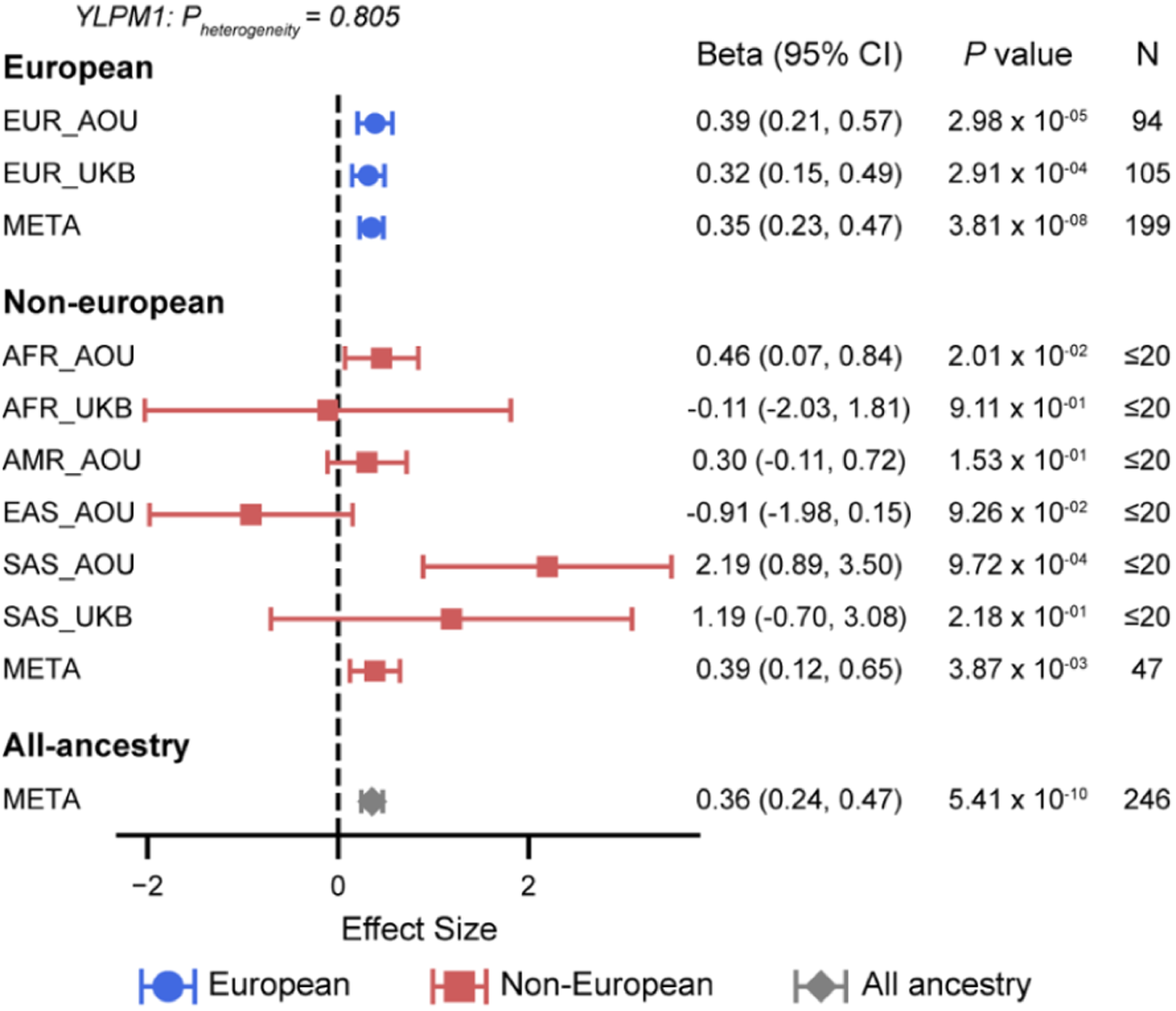
Effect of *YLPM1* PTVs on BMI across ancestries. 95% confidence intervals (CI) and significance values (*P* value) and number of heterozygous PTV carriers (N) of *YLPM1* gene on BMI across ancestries and biobanks. Three variant collapsing models were tested for the gene and the most deleterious mask that reached exome wide significance is shown here. Data for all genes contributing to changes in BMI are provided in **Supplementary Table 1**.

**Supplementary Fig. 2:**
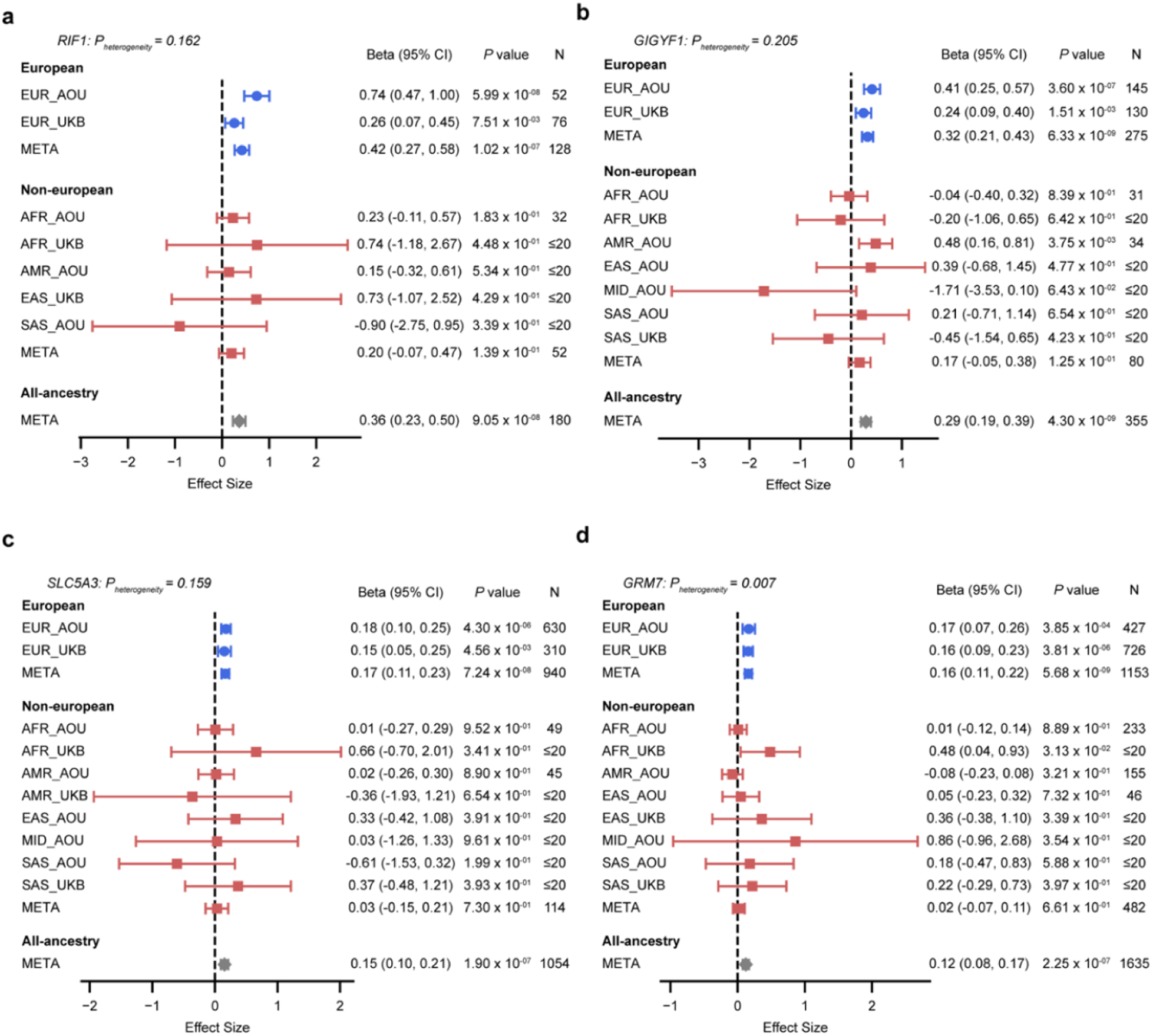
Effect of PTVs in discovered genes on BMI across ancestries. 95% confidence intervals (CI) and significance values (*P* value) and number of heterozygous PTV carriers (N) of (**a**) *RIF1*, (**b**) *GIGYF1*, (**c**) *SLC5A3*, and (**d**) *GRM7* genes on BMI across ancestries and biobanks. Three variant collapsing models were tested for the gene and the most deleterious mask that reached exome wide significance is shown here. Data for all genes contributing to changes in BMI are provided in **Supplementary Table 1**.

To gather supporting functional evidence for the discovered BMI associations, we extracted information from multiple sources including Genotype-Tissue Expression project (GTEx)^21^, International Mouse Phenotypic Consortium (IMPC)^22^, and Common Metabolic Diseases Knowledge Portal (CMDKP)^23^. The findings are summarized in **Table 2**. Notably, we observed *Ylpm1* heterozygous knockout mice to have significantly increased total body fat mass in males, as well as increased fasting circulating glucose level and decreased bone mineral content in a combined analysis of both males and females^22^. We also found “very strong” genetic support (Human Genetic Evidence, or HuGE scores^24^ ≥30) for *YLPM1* towards 12 cardiometabolic traits, including LDL cholesterol, total cholesterol, with obesity showing the highest evidence (HuGE score=133.61). Rare variant associations for obesity-associated traits have also been reported for *GIGYF1* and *RIF1* (**Table 2**). *For example*, large-scale sequencing analysis have linked *GIGYF1* and *SLC5A3* to increased waist-hip ratio adjusted for BMI and body fat percentage^25^. Additionally, copy number variations at the *GRM7* locus have been associated with severe early-onset obesity^26^, and a more recent study reported correlations between PTVs in *RIF1* and multiple obesity-related traits, including whole-body fat mass and hip circumference^10^.

Most obesity genes, including the five newly discovered BMI-associated genes, are primary expressed in the brain (**Table 2**) and are associated with nervous system disorders, such as neurodevelopmental disorders for *GRM7*^27^ and *GIGYF1*^28^ and bipolar disorder for *YLPM1*^16^. To ensure that the observed novel associations with BMI were not driven by the use of medications for psychiatric disorders (anti-psychotics and anti-depressants) which induce weight gain, we also accounted for medication intake and re-assessed gene burden associations for the five genes and found the results to be consistent with our original analysis (**Supplementary Table 6**).

**Table 2:** Supporting evidence for novel BMI-associated genes.

| Gene | Beta (BMI SD units) | Beta (BMI kg/m<sup>2</sup>) | Predicted Function | GTEx expression | IMPC Phenotype | CMDKP associations (with HuGE scores) |
| --- | --- | --- | --- | --- | --- | --- |
| *YLPM1* | 0.36 | 2.33 | RNA binding activity | Ubiquitously expressed, high in:<br>Brain (cerebellum)<br>Adipose (subcutaneous) | Increased total body fat (Male)<br>Higher fasting glucose (Combined) | Obesity (133.61)<br>LDL cholesterol (45)<br>Total cholesterol (45) |
| *RIF1* | 0.36 | 2.25 | DNA repair | Ubiquitously expressed, high in:<br>Brain (cerebellum)<br>Adipose (subcutaneous) | NA | Weight (348)<br>Body fat percentage (348) |
| *GIGYF1* | 0.29 | 1.46 | Insulin receptor signaling | Ubiquitously expressed, high in:<br>Brain (cerebellum)<br>Adipose (subcutaneous) | Not significant | Waist-hip ratio adj. BMI (105.86)<br>Waist circ. adj. BMI (60.16)<br>Total cholesterol (45). |
| *SLC5A3* | 0.15 | 0.92 | Inositol metabolic process | Artery (Aorta)<br>Kidney (Medulla) | NA | Not significant |
| *GRM7* | 0.12 | 0.72 | Glutamatergic neurotransmission | • Brain (Hypothalamus)<br>• Brain (Frontal Cortex) | Not significant | Not significant |

### Cardiometabolic profile of carriers of PTVs in BMI-associated genes

We examined whether PTV carriers in the BMI-associated genes were enriched across different obesity categories, namely, underweight/normal (BMI<25), overweight (25≤BMI<30), obese (30≤BMI<40), and severely obese (BMI≥40) in a combined analysis of all ancestries and biobanks. All eight known BMI-associated genes conferred increased risk for obese category (OR=1.33 to 2.74, *P*=2.08×10^-5^ to 1.22×10^-15^), while five (*MC4R*, *SLTM*, *UBR3*, *BSN*, and *BLTP1*) also conferred risk for severe obesity (OR=1.52 to 4.06, *P*=1.73×10^-5^ to 3.73×10^-12^) after multiple testing correction (**Supplementary Table 7,** *P*<9.61×10^-4^, Bonferroni correction for 13 genes and 4 obesity categories). Along the same lines with the known genes, PTV carriers in *YLPM1, RIF1, GIGYF1,* and *GRM7* were at an increased risk for both obese (OR=1.31 to 2.46, *P*=3.32×10^-7^ to 6.15×10^-10^) and severely obese (OR=1.18 to 2.26, *P*=3.80×10^-4^ to 2.33×10^-9^) categories (**Fig. 3a, Supplementary Table 7**). We did not observe significant enrichment for obese (OR=1.01, CI: 0.89, 1.15, *P=*0.87) or severely obese (OR=1.05, CI: 0.80, 1.35, *P=*0.69) clinical categories among *SLC5A3* carriers.

We also tested risks for 15 cardiometabolic comorbidities among PTV carriers of the obesity-risk genes compared to non-carriers (**Fig. 3b, Supplementary Table 8**). In addition to the known association of *BSN* (OR=2.63, 95% CI: 1.87, 3.64, *P=*5.61×10^-8^) and *GIGYF1* (OR=2.79, 95% CI: 2.18, 3.55, *P=*7.76×10^-15^) with type 2 diabetes (T2D)^12,29^, we also observed increased risk for hypertension (OR=1.75, 95% CI: 1.32, 2.33, *P=*1.03×10^-4^) and heart failure (OR=2.60, 95% CI: 1.59, 4.06, *P=*1.26×10^-4^) phenotypes among PTV carriers of *BSN*. Further, we found novel associations of *SLTM* and *SLC5A3* conferring increased risk for T2D (OR=2.19, 95% CI: 1.59, 2.98, *P=*1.73×10^-6^) and gastroesophageal reflux disease (OR=1.57, 95% CI: 1.36, 1.80, *P=*1.55×10^-9^), respectively.

To investigate the shared genetic architecture of obesity and its related traits, as well as to dissect the independent effect of the genes in the context of BMI towards increased comorbidity risk, we next performed structural equation modeling (SEM). SEM, represented by a path diagram, helps to understand the structure of relationships between independent and dependent variables in the context of other variables acting as mediators (**Supplementary Fig. 3**). With PTVs in a gene as an independent variable, BMI acting as a mediator, and the relevant comorbidity enriched in PTV carriers as the outcome, we observed distinct mechanisms through which the comorbidity risk increases (**Supplementary Table 9**). *For example*, we observed BMI acting as a partial mediator for T2D risk in *BSN*, *GIGYF1*, and *SLTM* carriers where the direct effect of the genes (**Fig. 3c, d and e**, *BSN*: β=0.34, CI: 0.16, 0.53, *P=*3.31×10^-4^, *GIGYF1*: β=0.46, CI: 0.33, 0.60, *P=*2.53×10^-11^, *SLTM*: β=0.24, CI: 0.07, 0.41, *P=*5.51×10^-3^) was higher than the indirect effect through BMI (*BSN*: β=0.13, CI: 0.09, 0.16, *P=*4.07×10^-10^, *GIGYF1*: β=0.09, CI: 0.06, 0.12, *P=*1.23×10^-9^, *SLTM*: β=0.15, CI: 0.12, 0.19, *P=*3.40×10^-17^). On the other hand, we observed no mediation through BMI (β=0.006, CI: −0.001, 0.014, *P=*0.11) but a significant direct effect of *SLC5A3* on increased risk for gastroesophageal reflux disease (β=0.19, CI: 0.10, 0.27, *P=*1.37×10^-5^) (**Fig. 3f**).

**Supplementary Fig. 3:**
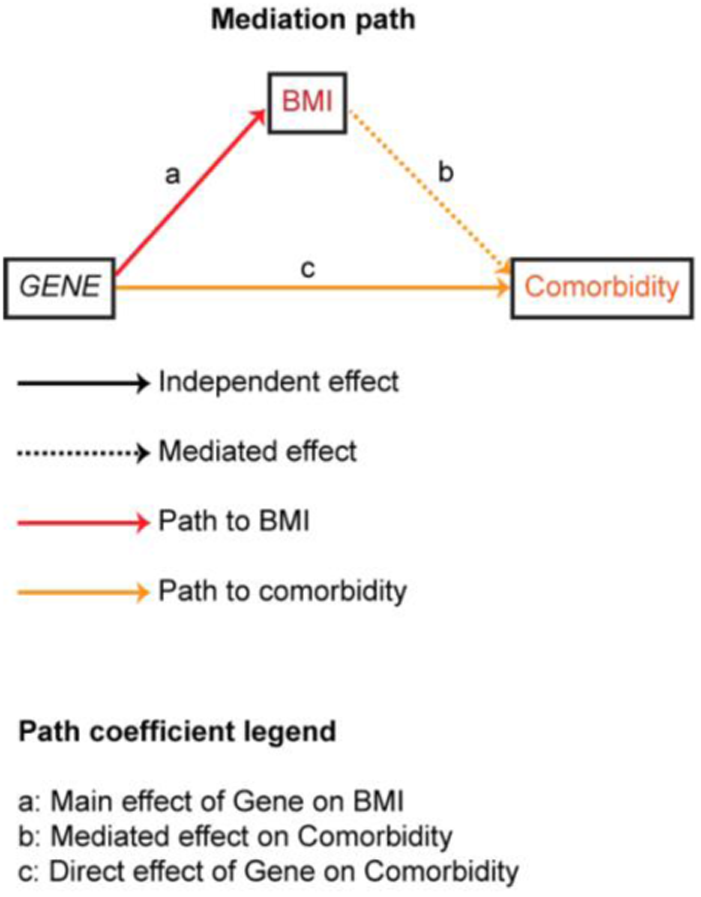
Structural Equation Modeling schematic. Path diagram demonstrating the risk conferring effects of BMI associated genes on comorbidities obtained using mediation analysis.

**Fig. 3:**
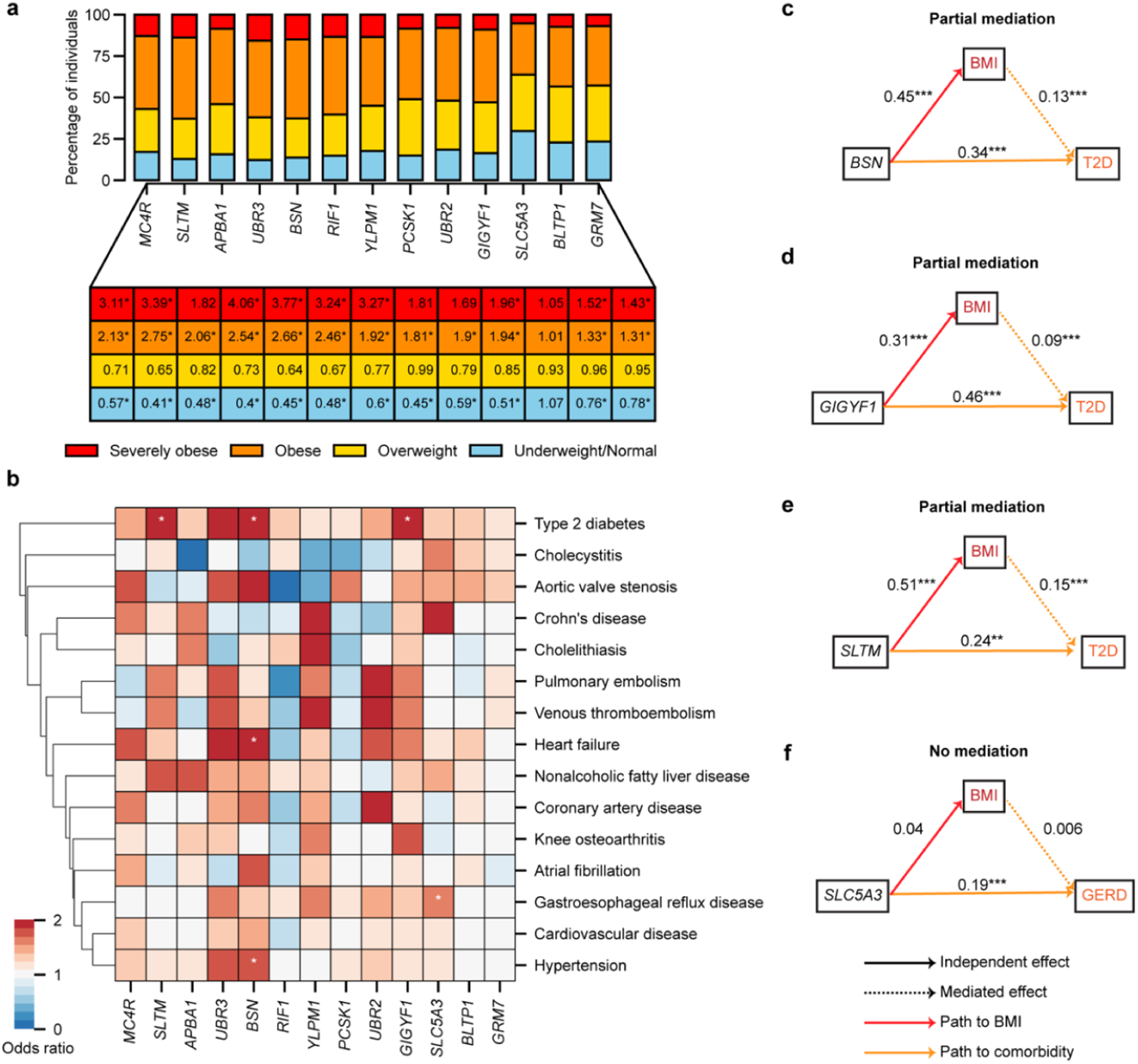
Enrichment of obesity and cardiometabolic comorbidities in PTV carriers of the discovered BMI associated genes. (**a**) *Top*: Proportion of PTV carriers within the obesity clinical categories (underweight or normal, overweight, obese, and severely obese) for BMI associated genes. *Bottom*: Odds ratio of obesity clinical categories in carriers compared to non-carriers. \**P*<9.61×10^-4^ (Bonferroni corrected for 13 genes and four categories). Extended data with the odds ratios, 95% confidence intervals and exact *P* values are available in **Supplementary Table 7.** (**b**) Odds ratio for obesity-related comorbidities in PTV carriers of risk or protective genes compared to non-carriers. \**P*<2.56×10^-4^ (Bonferroni corrected for 13 genes and 15 comorbidities). Extended data with the odds ratios, 95% confidence intervals and exact *P* values are available in **Supplementary Table 8**. Structural equation models depicting the path through which the gene affects increased comorbidity risk with BMI acting as a (**c**) partial mediator for *BSN* and type 2 diabetes (T2D), (**d**) partial mediator for *GIGYF1* and T2D, (**e**) partial mediator for *SLTM* and T2D, and (**f**) not mediating gastroesophageal reflux disease (GERD) in *SLC5A3* carriers. \*\**P*<0.01, \*\*\**P*<0.001. Extended data with statistics for the entire model are available in **Supplementary Table 9.**

Using a phenome-wide study, we also identified additional associations for the five discovered genes (**Supplementary Tables 10**). *For example*, *YLPM1* PTV carrier status was associated with “mental disorders” such as altered mental status (**Fig. 4a**, β=1.33, CI: 0.79, 1.87, *P=*1.41×10^-6^) and “digestive disorders” such as cholelithiasis (β=0.94, CI: 0.50, 1.38, *P=*2.48×10^-5^). In addition, *GIGYF1* showed increased risk of hypothyroidism (β=1.17, CI: 0.88, 1.46, *P=*1.98×10^-15^) and chronic renal failure (β=1.01, CI: 0.61, 1.41, *P=*9.98×10^-7^) along with the already identified association with T2D (**Fig. 4b**).

**Fig. 4:**
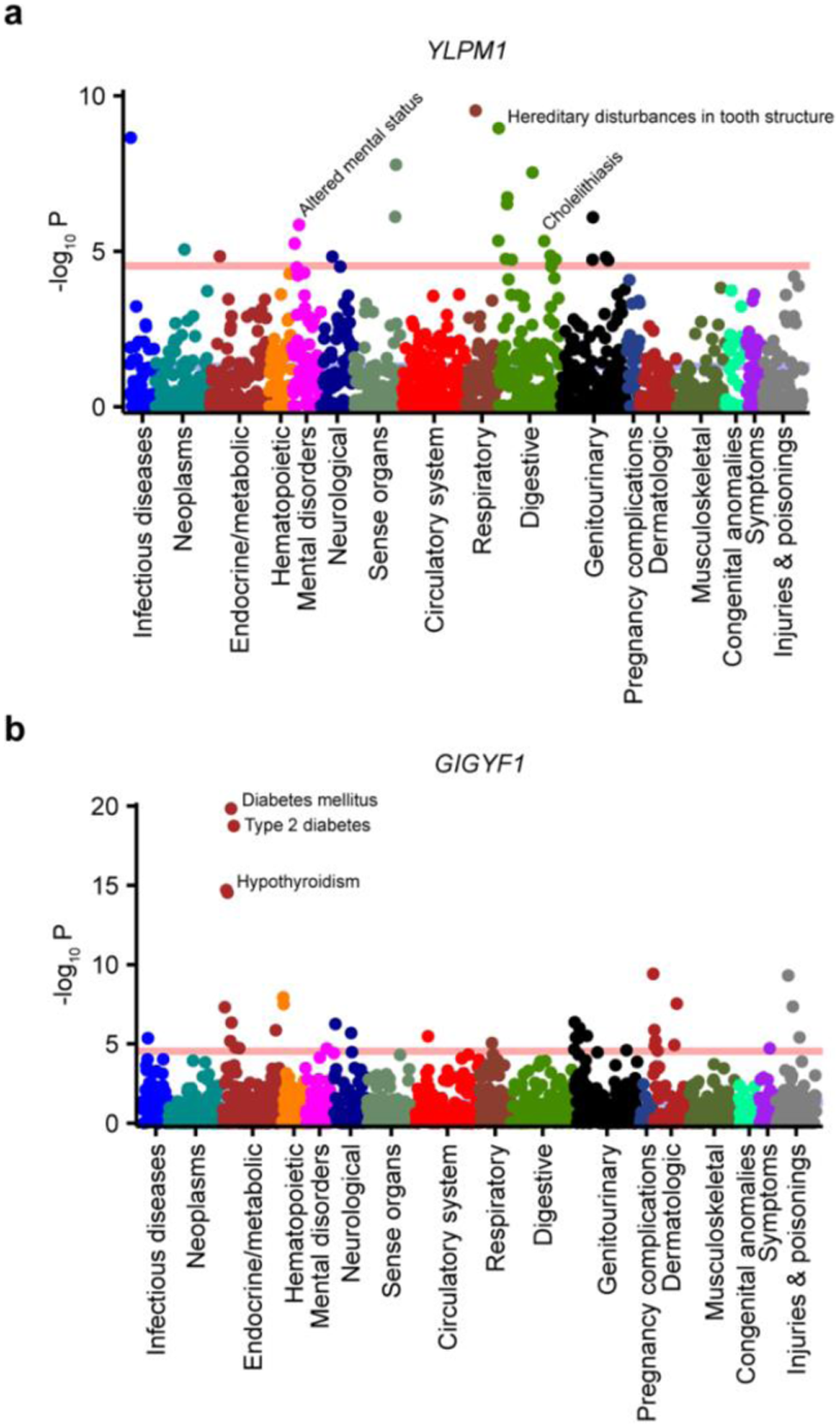
Phenome-wide characterization of BMI-associated genes. PheWAS results of **(a)** *YLPM1* and **(b)** *GIGYF1* demonstrating the associated phenotypes clustered by their broader categories based on phecodes definitions. Pink line denotes the multiple testing significance threshold (Bonferroni cutoff). Significant PheWAS results for all five discovered genes are available in **Supplementary Table 10**.

### Influence of PGS on obesity risk conferred by rare variants

Previous studies investigating the effect of polygenic burden on obesity risk in rare variant carriers have suggested an additive influence of common variants in *MC4R* and non-additive in *BSN* PTV carriers^9,12^. We used linear regression to test multiplicative interactions between PGS and each disrupted gene towards changes in BMI among Europeans. Our meta-analysis of both UKB and AoU cohorts suggest polygenic burden affects BMI and obesity penetrance in an additive manner, including in *BSN* carriers (**Supplementary Fig. 4a, Supplementary Table 11**). Among PTV carriers of all obesity genes in UKB, there was a steady rise in obesity prevalence from 22% among individuals in the lowest PGS quintile to 48% among individuals in the highest quintile (**Supplementary Fig. 4b**). Similarly, in AoU, obesity prevalence rose from 38% to 54% from the lowest to the highest quintile among PTV carriers of obesity genes (**Supplementary Fig. 4b**). The difference in mean BMI between PTV carriers in the highest PGS quintile and those in the lowest quintile was 3.88 and 2.63 kg/m^2^ in UKB and AoU, respectively (**Supplementary Fig. 4b, Supplementary Table 12**).

**Supplementary Fig. 4:**
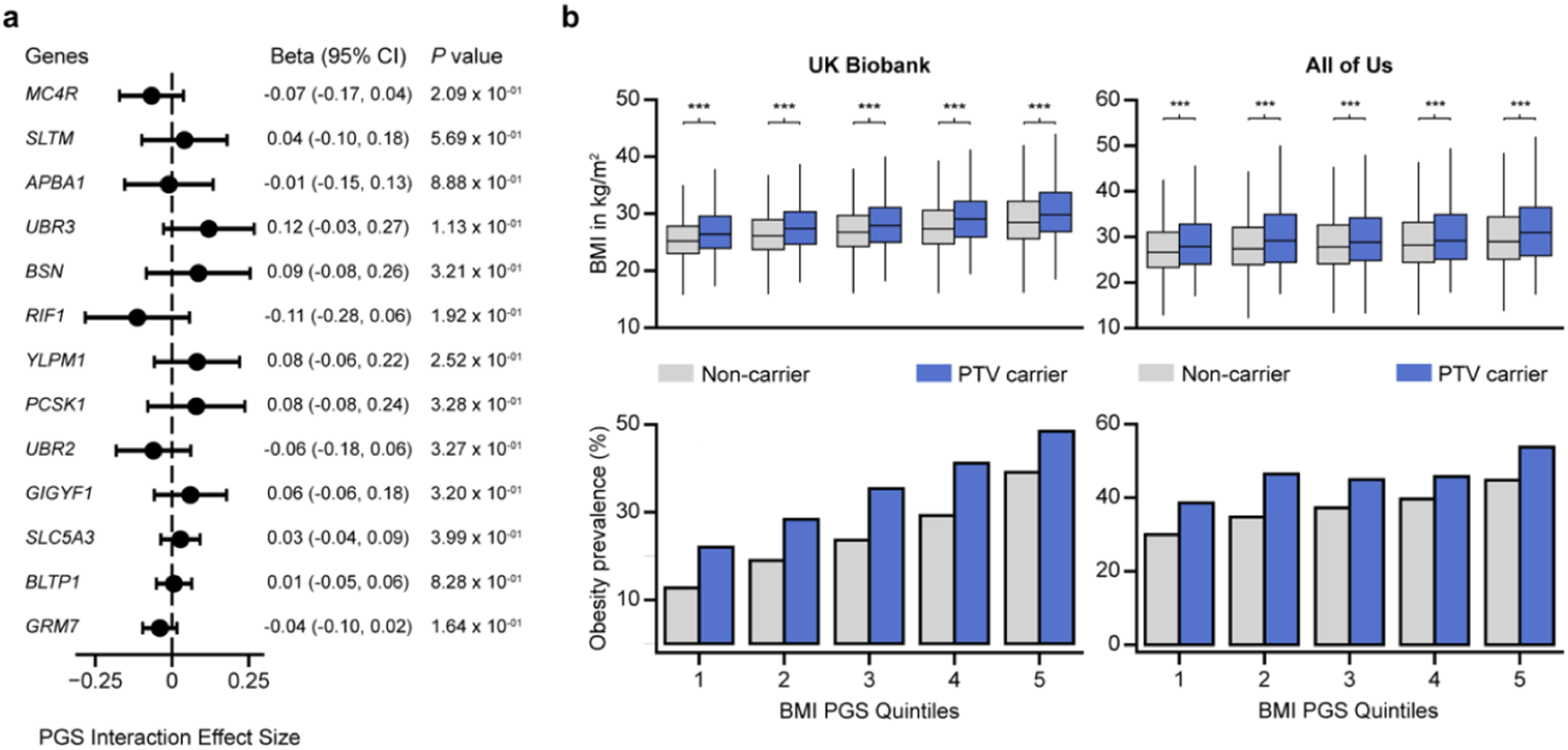
Effect of obesity-associated genes modulated by polygenic risk. (**a**) Interaction model coefficient plot of combined risk of PTVs in the discovered genes and PGS on BMI. Data with exact *P* values and other associated statistics are available in **Supplementary Table 11. (b**) BMI distribution (*top*) and obesity prevalence (*bottom*) across PGS quintiles of PTV carriers (blue) compared to non-carriers (grey) of BMI-associated genes in UKB (*left*) and AoU (*right*) cohorts. Data with exact *P* values and other associated statistics are available in **Supplementary Table 12**.

### Effect of obesity-associated genes on plasma protein levels

We next evaluated the impact of PTVs in the obesity-associated genes on the levels of 2,923 plasma proteins available for approximately 50,000 individuals in UKB. We detected significant changes in plasma protein levels with PTV carrier status for four genes (**Supplementary Fig. 5, Supplementary Table 13**). *For example*, we identified *SLTM* PTV carriers had increased LECT2 (β=1.27, CI: 0.69, 1.84, *P*=1.48×10^-5^) protein levels. LECT2 is a hepatokine that has been previously linked to obesity^30^, and we found significant association of LECT2 levels with BMI in UKB as well (β=0.21, CI: 0.20, 0.22, *P*<1×10^-100^, **Fig. 5, Supplementary Table 14**). Additionally, *GIGYF1* PTV carriers had decreased ODAM (β=-0.76, CI: −1.09, −0.42, *P*<1.10×10^-5^) and NCAN (β=-0.67, CI: −0.97, −0.37, *P*<1.41×10^-5^) protein levels, while *BLTP1* PTV carriers had increased CD164 (β=0.51, CI: 0.30, 0.73, *P*<2.24×10^-6^) and TNFSF12 (β=0.48, CI: 0.27, 0.69, *P*<5.76×10^-6^) protein levels, and we further found the levels of all these proteins to also be significantly associated with BMI (**Fig. 5, Supplementary Table 14**). Our results highlight the potential downstream protein targets through which obesity-associated genes might act to cause the phenotype.

**Fig. 5:**
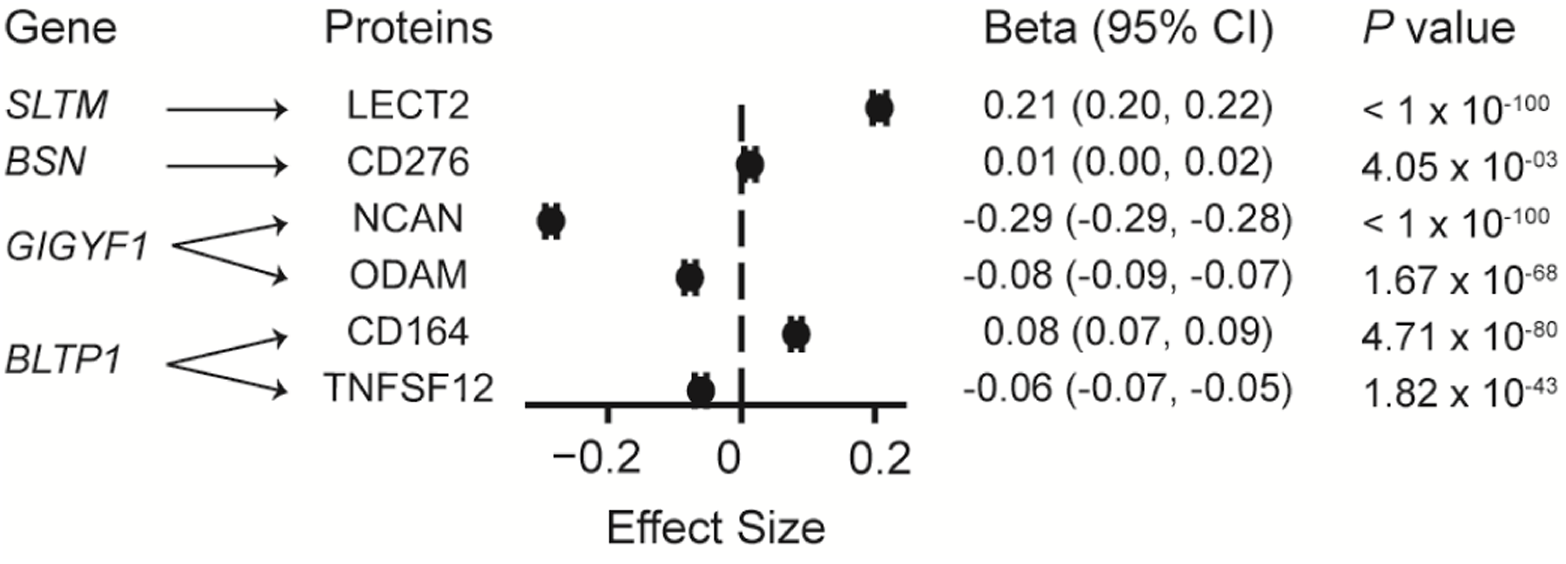
Relationship between plasma protein levels and BMI. Model coefficients, with 95 % CI and *P* value, of a linear model constructed using plasma protein levels as independent variable and BMI as the dependent variable adjusting for covariates (see **Methods**). Only proteins where PTV carrier status of BMI associated genes were significantly associated with their expression level were used to create protein models of BMI. Data with exact *P* values, model coefficients and 95% confidence interval are available in **Supplementary Table 14.**

**Supplementary Fig. 5:**
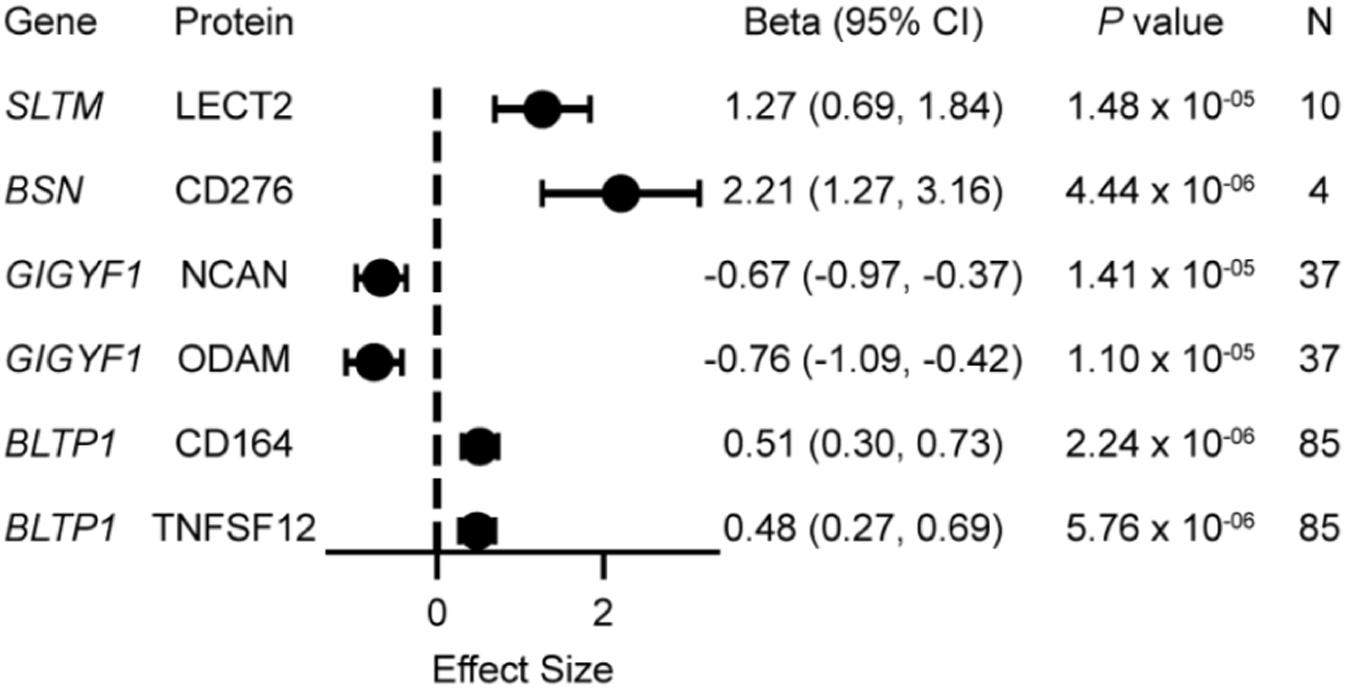
Impact on plasma protein of BMI-associated genes. Model coefficients, with 95 % CI and *P* value, of a linear model constructed using PTV carrier status of BMI associated genes as independent variable and plasma protein expression value as the dependent variable adjusting for covariates (see **Methods**). Only proteins where PTV carrier status were significantly associated with expression level (*P*<1.7×10^-5^, Bonferroni correction for 2923 proteins) are shown. Data with exact *P* values, model coefficients and 95% confidence interval are available in **Supplementary Table 13.**

### Effect of previously identified obesity genes on BMI across ancestries and biobanks

We next examined the strength of association of previously identified obesity-associated genes that did not pass statistical significance in our study. Of the four genes with pLoF variants reported by Akbari and colleagues, three genes, including *ROBO1*, *ANO4*, and *GPR75* showed suggestive significance in our dataset (*P<*10^-6^, **Supplementary Fig. 6a, Supplementary Table 15**), reinforcing the potential role of those genes in obesity^9^. Akbari and colleagues also identified nine additional BMI-associated genes based on both pLoF and deleterious missense variants, but due to substantial differences in the deleterious missense prediction methods used in the two studies (see **Methods**) a direct comparison could not be performed. By conducting a sex-stratified study across ancestries, we also examined the effect of three female-specific obesity genes (*DIDO1*, *PTPRG*, and *SLC12A5*) identified by Kaisinger and colleagues^19^. While we found similar trends among Europeans, where females carriers showed increased BMI and males remained largely unaffected, these patterns did not reach exome-wide significance across ancestries (**Supplementary Fig. 6b, Supplementary Table 16**).

**Supplementary Fig. 6:**
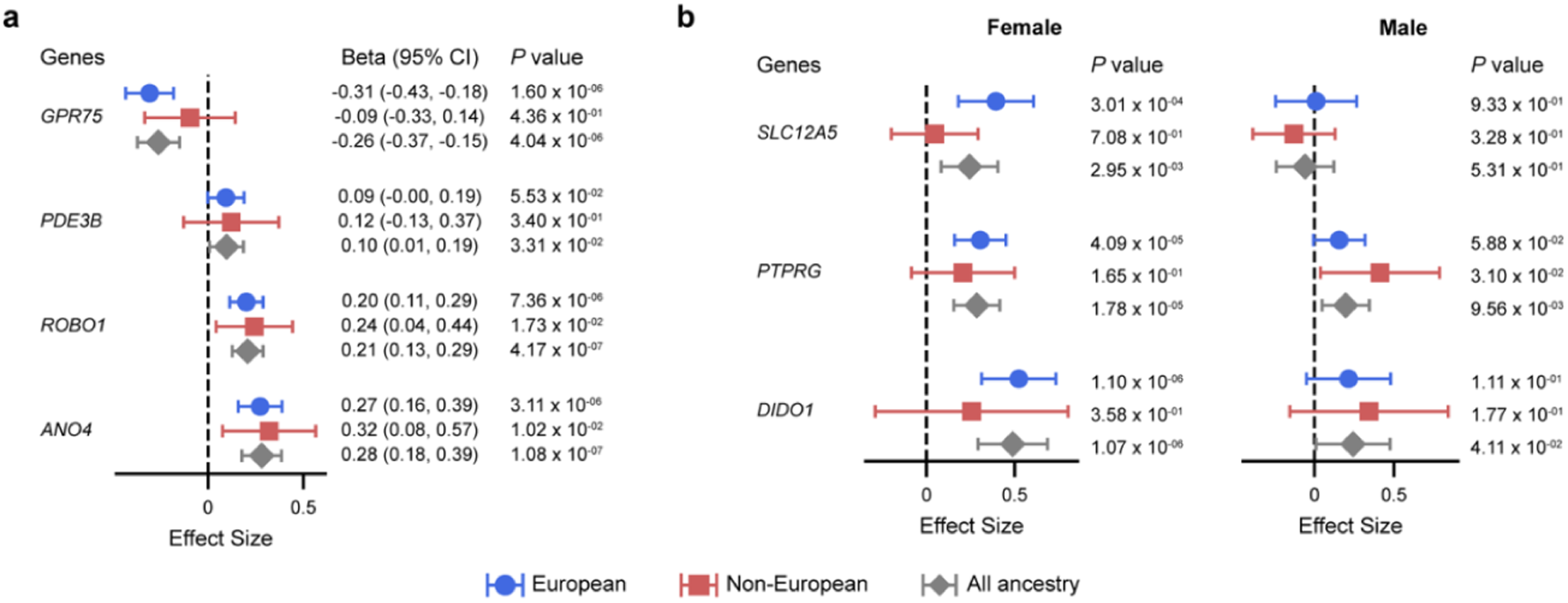
Meta analysis results of previously identified BMI associated genes. **(a)** Effect sizes along with 95% confidence intervals (CI) and significance values (*P* value) in European, non-European, and combined meta-analysis for previously identified obesity genes with pLoF variants on BMI in our study. Data and additional statistics are provided in **Supplementary Table 15**. (**b**) Effect sizes on BMI along with 95% confidence intervals (CI) and significance values (*P* value) in European, non-European, and combined meta-analysis for previously identified female-specific obesity genes with pLoF variants in females (left) and males (right) in our study. Data and additional statistics are provided in **Supplementary Table 16**.

## DISCUSSION

Here, we discovered five novel genes that associated with increased BMI, four of which (*YLPM1*, *RIF1*, *GIGYF1*, and *GRM7*) also associated with obesity and severe obesity in a multi-ancestry analysis. The effect sizes of *YLPM1* and *RIF1* were comparable to canonical obesity genes such as *MC4R* and *BSN*. The associations with BMI discovered in the European cohort was confirmed in a non-European cohort and were also supported by additional evidence, such as increased fat body mass in previously phenotyped mouse KO model for *Ylpm1*. We also examined the impact of known obesity-related genes across multiple ancestries, leveraging additional AoU data that prioritized the recruitment of individuals from less studied genetic backgrounds, including African and Native American populations. While some genes such as *YLPM1*, *MC4R*, and *SLTM* showed consistent effects across ancestries, others such as *GRM7* and *APBA1* showed a significant European bias, which could impact their potential as therapeutic targets for obesity.

In addition to obesity, a phenome-wide association study of PTV carriers of the novel genes revealed further associations with obesity-related comorbidities, for example, *YLPM1* with cholelithiasis and *GIGYF1* with T2D and hypothyroidism. In addition, we also observed association of *YLPM1* with “mental disorders” such as altered mental status. While prior studies have reported associations between *YLPM1* and psychiatric disorders^16^, our analysis demonstrates that *YLPM1* influences metabolic disorders as well, highlighting its pleiotropic effect on multiple phenotypes. This is further supported by the ubiquitous expression of *YLPM1* across multiple tissues and its potential role in transcriptional regulation. Moreover, obesity-associated genes, including the novel ones identified in this study (**Table 2**), have been implicated in neuronal processes, including appetite regulation, neurodevelopment, and neurogenesis^4,6,31^. Nevertheless, further functional characterization of *YLPM1* is essential to elucidate the biological mechanisms through which it might influence obesity and related disorders.

Although additive effects are the most common mechanism underlying the combined influence of high-effect rare variants and low-effect common variants on obesity susceptibility, a recent study suggested a potential non-additive risk increase driven by PGS in *BSN* PTV carriers^12^. This observation, however, was limited to a homogeneous population of White British individuals from a single cohort. In contrast, our analysis of European individuals across multiple cohorts does not provide sufficient evidence to support non-additive interactions between common variants and rare PTVs for any of the discovered genes. However, we observed a substantial effect of the combined influence of both risk factors on obesity penetrance across cohorts.

Along with non-additive interactions between obesity-associated variants, protective obesity genes, often driven by variants with lower frequencies and smaller effect sizes, have typically been more challenging to identify than risk genes. Notable examples such as *GPR75*, *GPR151*, and *GIPR* have only recently emerged through large-scale rare variant association studies^6,9,16^. Despite leveraging datasets from multiple cohorts across ancestries, our study remains underpowered to detect protective effects, as evidenced by *GPR75* reaching only a suggestive threshold of significance. Among genes with negative effect sizes in our combined cross-ancestry analysis, *PLCB4* and *LMX1B* displayed effect sizes, PTV carrier counts, and *P* values comparable to *GPR75*. While nearby common variant signals associated with BMI have been reported previously for these two genes, rare variant associations did not reach exome-wide significance in our study, suggesting that larger datasets will be required to confirm their associations with obesity.

So far, gene discoveries related to obesity have predominantly focused on individuals of European ancestry, largely due to limited sample sizes from non-European populations. Despite commendable efforts by the AoU program to build an ancestrally varying biobank, our results suggest that current sample sizes remain underpowered to enable ancestry-specific gene discoveries in non-European populations. For instance, no genes reached exome-wide significance in any non-European ancestry group, whereas five genes crossed the threshold in UKB Europeans and another five in AoU Europeans, underscoring the need for larger sample sizes to enable robust ancestry-specific gene discoveries beyond European populations. Nevertheless, *YLPM1*, *MC4R*, and *SLTM* showed consistent effects on BMI across multiple ancestries and cohorts highlighting the generalizability of these associations across various genetic backgrounds, environmental exposures, and ascertainments. Overall, using a multi-ancestry approach, we provide a more comprehensive view of obesity genetics, identifying novel genes, refining the effects of known genes, and demonstrating the importance of including heterogenous populations in genetic studies.

## METHODS

### SNP array quality control and filtering

We accessed SNP array data of 488,377 individuals released by UKB and available in plink format in UKB Research Analysis Platform (RAP). First, we lifted over the SNPs from GRCh37 to GRCh38 format using Picard’s LiftoverVCF tool^32^. We then applied the quality control filters on the lifted over SNP and retained SNPs with minor allele frequency (MAF) >0.01, minor allele count (MAC) >100, missing call rate <0.1, and Hardy-Weinberg equilibrium exact test *P* value (HWE *P*) >1×10^-15^. The filtered SNPs were further LD pruned with an *r*^2^ threshold of 0.9, 1000 variants window size and 100 variant window slides using plink v2^33^. Samples with missing call rate >0.1 were also removed.

Similarly, for AoU we accessed array data of 447,278 individuals and applied the same filters using plink v2 in the AoU Researcher Workbench.

### Variant quality control, filtering, and annotation

We analyzed whole exome sequencing data of 469,835 individuals from the UKB cohort, available as multi-sample project variant call format (pVCF) files in the UKB Research Analysis Platform (RAP). The sequencing method and preparation of the pVCF files have been previously described^34^. Using the Hail^35^ platform on DNANexus, we *first* filtered low quality genotypes and variant calls. Genotype calls with genotype quality (GQ) >=20, depth (DP) >=10 (5 for haploid genotypes on sex chromosomes), and allele balance (AB) >= 0.2 and <=0.8 (for heterozygous genotypes only) were retained following gnomAD guidelines^36^. Variants with call rate >90%, HWE *P* >1×10^-15^, not within Ensembl low-complexity regions and not monomorphic, were retained. *Second*, we performed sample quality control using a set of high-quality autosomal and X-chromosome variants (see **Supplementary Notes**). Samples failing quality control parameters, such as missing microarray data, duplicated samples, sex discordance, <90% exome variant call rate, or significantly deviated from ancestry residualized genetic quality-control metrics, were flagged (**Supplementary Notes**). *Third*, we extracted the quality controlled exome variants retaining rare variants (MAF <0.01%) present within the autosomes and annotated them using variant effect predictor^37^ (VEP v109) and dbNSFP v4^38^ to identify the predicted variant effects on gene transcripts. We categorized each variant based on their most deleterious functional impact on the transcript into three categories, from the most harmful to the least harmful, as follows: (1) predicted loss of function or “pLoF”, which included frameshift, stop gained, splice acceptor, and splice donor VEP annotations, (2) “Missense strict”, which included missense variants predicted to be deleterious by nine deleteriousness prediction tools (SIFT^39^, LRT^40^, FATHMM^41^, PROVEAN^42^, MetaSVM^43^, MetaLR^43^, PrimateAI^44^, DEOGEN2^45^, and MutationAssessor^46^) available through dbNSFP database, and (3) “Missense lenient”, which included missense variants predicted to be deleterious by at least seven out of the nine deleteriousness prediction tools. After filtering out the flagged samples from sample quality control step, we collected the sample ids of both heterozygous and homozygous samples carrying the retained variants. *Finally*, we added gnomad exome MAF annotations of five superpopulations (AFR, AMR, EAS, SAS, EUR) to the retained variants using VEP v109 and Hail. For each variant we assigned an alternate allele frequency (AAF) based on the maximum of the MAF observed in the UKB cohort and the five gnomad annotated superpopulations. All UKB analyses were performed in the DNANexus UKB RAP.

We also analyzed whole genome sequencing data of 414,830 individuals in the AoU cohort. Variant call files for regions overlapping with the exomes were available as a Hail matrix table in the AoU portal. The AoU team has centrally performed extensive variant and sample quality control filtering using genomic data before its release and made all quality control information available through the research portal. In addition to the centrally performed quality control, we further filtered genotype and variant calls based on gnomad guidelines as described for UKB variant quality control. For sample quality control, we flagged all the samples that failed AoU quality control parameters using AoU provided quality control files. In addition, we removed possible duplicate samples based on available kinship scores >0.354 and those with sex at birth not reported as Male or Female. After filtering the variants for intracohort frequency (<0.001), we annotated the variants using Nirvana^47^, available in the AoU research platform as a Hail table, and determined the functional impact of each variant and the heterozygous or homozygous samples for the variants using the same criteria as defined for the UKB data. Additionally, using gnomad genome MAF annotations for the five superpopulations available through NIRVANA, we assigned AAF to each variant based on the maximum of the MAF observed in the UKB cohort and the five gnomad annotated superpopulations. All AoU analyses were performed in the Researcher Workbench of the AoU portal.

### Ancestry inference

For inferring ancestries of each sample in UKB, we followed the same protocol as previously used to predict ancestry in AoU^15^. In brief, we extracted samples with labelled ancestries from the Human Genome Diversity Project and the 1000 Genome project consisting of 4,151 individuals^48^. After filtering related samples and those who failed gnomad quality control filters, 3,224 samples were retained to use as reference samples. We then identified overlapping SNPs between reference and UKB array and further filtered the overlapped sites to retain autosomal variants with MAF >0.1%, call rate >99% and LD pruned variants with a cutoff r2 of 0.1 to obtain maximal subset of uncorrelated variants within a 1-Mbp window. We next generated the first 16 principal components (PC) of the reference sample using “hwe normalized pca” function in Hail. We then projected the genotypes of the UKB samples to the PC space of the reference samples using “pc project” function in Hail. The 16 PCs were used as feature vectors for the reference samples to train a random forest model using scikit learn with six gnomad assigned continental ancestries (African (afr); South Asian (sas); East Asian (eas); Middle Eastern (mid); Latino/admixed American (amr); European (eur), composed of Finnish (fin) and Non-Finnish European (nfe) as labels. The trained model was used to predict probabilities of the six ancestries among the UKB samples. Samples with predicted ancestral probability >75% were assigned that ancestry while individuals with <75% probability for all ancestries were assigned as “other”. We assessed the concordance of the predicted ancestries with self-reported ethnicities, and report our findings in the **Supplementary Note**.

In AoU, we extracted previously calculated genetic ancestry^15^ of each sample from auxiliary files available through AoU researcher workbench.

### Phenotype, obesogenic lifestyle, polygenic risk analyses

***For the UKB cohort***, BMI (Data Field 21001), genetic sex (Data Field 22001), age (Data Field 21003), ethnic background (Data Field 21000), BMI PGS (Data Field 26216), genetic kinship (Data Field 22021), and the top 10 genetic principal components (PCs; Data Field 22009) on 502,368 individuals in UKB were accessed and preprocessed through the UKB RAP. Numerical fields such as BMI and age, measured across multiple visits, were averaged. International Classification of Disease (ICD-10) 10^th^ revision summary diagnosis codes corresponding to each individual were also extracted from the Hospital Episode Statistics (HES) data available in the UKB RAP. For each ICD code, we obtained the samples who were assigned that particular code as well as any lower-level ICD codes which fell under the hierarchy of the original code based on the hierarchical ICD tree. We next obtained the specific ICD codes for obesity associated comorbidities based on previous BMI related studies and annotated individuals as carrying a comorbidity if they were diagnosed with the corresponding ICD codes. A list of the comorbidities and the ICD codes used to categorize an individual as carrying the comorbidity is provided in **Supplementary Table 17**. Finally, we filtered samples without exome data, those who failed any of the genetic data quality control filters, those without BMI information, those without ancestry predictions or ancestry predicted as “oth” and with 10 or more third-degree relatives from our analysis based on previously published kinship estimates^17^, resulting in a final cohort of 454,645 individuals with exome, phenotype, and covariate data.

***For the AoU cohort***, we obtained BMI, age, genetic sex, 10 genetic PCs, and sample IDs of related individuals from the AoU Workbench. We calculated the age based on the date of birth and the BMI measurement data concepts (“concept” defined here as a collection of similar data stored together) available in AoU. We used the “sex at birth” concept, available in AoU, to denote the genetic sex of an individual. The genetic sex of all individuals with survey sex listed as Male and Female in AoU were concordant as per centrally performed quality control statistics. Therefore, we retained individuals with “sex at birth” field listed as either “Male” or “Female” as well as those with BMI values between 12 and 75 kg/m^2^ to ensure consistency between UKB and AoU. Additionally, we filtered individuals who failed genetic quality control parameters and those without ancestry predictions or with ancestry predicted as “oth”, resulting in a final cohort of 384,465 individuals.

We calculated ***PGS for BMI*** in individuals in the AoU cohort by using the GWAS summary statistics from a previous study^4^. Duplicate and ambiguous single nucleotide polymorphism (SNPs) were filtered from the summary statistics. Genotype data of the AoU cohort filtered for common variants (MAF>0.01) was obtained as a Hail matrix table. We further removed variants with HWE *P* value <1×10^-6^ and retained variants that overlapped between summary statistics data and the AoU genotyped data with a call rate >0.9. We then used the predetermined effect sizes of the SNPs as Bayesian priors, weighed them based on the AAF observed in individuals, and summed the weighted effect sizes to obtain individual-specific polygenic risk for BMI. All calculations were conducted using Hail, available in the AoU Researcher Workbench. ICD-10 diagnosis codes corresponding to each individual in the AoU cohort was extracted using the researcher workbench. We next obtained the specific ICD codes for obesity associated comorbidities based on previous BMI related studies and annotated individuals as carrying a comorbidity if they were diagnosed with the corresponding ICD codes as performed in UKB.

The prepared phenotype files with BMI, covariates, BMI PGS and obesity-associated comorbidities from both cohorts were separated into six ancestry-specific phenotype files. BMI was rank inverse-normal transformed for each ancestry, grouped by sex of individuals. Finally, we binarized genetic sex field, assigning value of 1 to females and 0 to males, and calculated age^2^ and age × sex covariate terms.

### Gene burden association test using REGENIE

We used REGENIE v3.3^18^ to conduct gene burden association tests. Similar to most whole genome regression tools, REGENIE operates in two steps. *First*, it uses SNP data, preferably a genotyped array of SNPs, from across the genome to fit a null model that estimates a polygenic score for the trait to be tested (i.e., normalized BMI in our study). This step accounts for population structure and relatedness between samples. For both UKB and AoU cohorts, we used the already prepared and quality-controlled SNP array files. In the *second* step, REGENIE calculates associations between the genetic variants of interest and the trait after considering the null model, calculated in step 1, and other user-defined covariates. In our study, we used age, age^2^, sex, age × sex and first ten genetic PCs obtained from both UKB and AoU cohorts as additional covariates. In UKB, exome release batch was added as an additional covariate.

REGENIE is capable of collapsing variants to a gene-level with user defined masks and annotation files apart from the usual variant data as input before running association tests. We supplied our already annotated variant file based on their impact on gene transcripts and defined three variant masks to collapse variants on a gene level, namely, (a) “pLoF” variants only, (b) “pLoF” and “Missense strict” variants, and (c) “pLoF”, “Missense strict” and “Missense lenient” variants. Additionally, the AAF of the variants to be considered within a gene mask could also be specified. We provided the previously assigned AAF of the variants calculated based on the maximum of cohort frequency and gnomad annotated superpopulation frequencies as input. We performed independent gene-based rare variant association tests for all the six continental ancestries across both biobanks and generated statistics for all individual gene-mask pairs considering variants with a maximum AAF of 0.001.

### Meta analysis and ancestral heterogeneity calculation

Results from the individual ancestry study in each biobank were pooled into the three meta statistics (a) European: combined eur ancestry statistics from AoU and UKB, (b) Non-European: combined afr, amr, eas, sas, and mid ancestry statistics from AoU and UKB, and (c) All ancestry: combined all six ancestry statistics from AoU and UKB independent tests using an inverse variance-weighted (IVW) fixed-effects model. The meta-analysis calculation was implemented using the python package SciPy and NumPy packages with in-house generated scripts. Each gene-variant mask was associated with meta statistics for three meta populations. Any gene-variant mask that passed the Bonferroni multiple testing correction threshold of 8.34×10^-7^, accounting for 20,000 genes and three variant collapsing models, in the “European meta” ***and*** in the “All ancestry meta” population was considered to be significantly associated with BMI across ancestries. We also used the meta-analysis tool METAL to calculate combined statistics for the three meta studies while controlling for genomic inflation rates for the independent rare variant burden tests^20^.

To assess heterogeneity in effect sizes between European and non-European ancestries, we applied Cochran’s Q test, a standard approach for assessing between-group variation in meta-analyses. Effect sizes and their corresponding standard errors from each ancestry group were used to compute the Q statistic, which quantifies deviation from the weighted mean effect size.

The weights were determined based on the inverse variance of each effect size estimate. The p-value for heterogeneity was obtained by comparing the Q statistic to a chi-squared distribution with one degree of freedom. All statistical analyses were conducted in Python using NumPy and SciPy.

### Sensitivity analysis

For the gene burden associations of the novel five genes identified in this study that crossed exome-wide significance in our cross-ancestry analysis, we performed LOVO analysis which creates burden masks by iteratively excluding a single variant and recalculating the gene burden association statistics (CITE). If a single variant contributed to the significant association of the gene, then the burden study after leaving the variant will result in a substantially decreased association statistic for that mask, enabling the identification of such variant thorough LOVO analysis.

We also tested for the effects of shadow effect of nearby common variants within a 2-Mbp locus of the start coordinates of the genes which were significant in the independent gene burden test. Any common variant that falls within the locus and show suggestive association with BMI (P <0.01), based on a previous study^4^, was included as additional covariates in a reanalysis of the gene-burden association for the five novel genes identified in our study.

Additionally, we reanalyzed effect of the five novel genes on BMI by accounting for medication intake (antipsychotic and antidepressants) as an additional covariate in the gene-burden association tests. *In the UKB cohort*, we extracted the medications consumed data by each individual (Data Field 20003) and categorized the medications based on Anatomical Therapeutic Chemical (ATC) Classification System codes using mappings prepared by a previous study^49^. Subsequently, we categorized the individuals in the cohort based on whether or not they consumed medications classified as ATC codes N06A for anti-depressants and N05A for anti-psychotics. *In the AoU cohort*, medications were already mapped to their ATC codes. We extracted samples who consumed medications that fell in either of the two categories, N06A or N05A. Finally, we used the defined medication consumption field as an additional covariate in the gene-burden association tests to recalculate effect sizes of the novel genes on BMI.

### Odds-ratio calculation for obesity clinical categories and obesity-related disorders

To calculate risk across the obesity clinical categories among carriers of PTVs in the discovered genes compared to non-carriers, we first categorized each individual into their respective clinical category based on their BMI values: underweight or normal (<25), overweight (25 to 30), obese (30 to 40) and severely obese (>40). We then created 2×2 contingency tables using their carrier status as the first variable and whether they belong to the respective obesity category as opposed to any other category as the second variable. Finally, we calculated the conditional odds ratio and the significance value using a two-sided Fisher’s exact test.

To calculate risk for obesity related disorders, we obtained the specific ICD codes for comorbidities frequently associated with obesity, based on previous BMI related studies^1,3^, and calculated the odds of PTV carriers diagnosed with the comorbidity compared to non-carriers using Fisher’s exact test. A list of the comorbidities and the ICD codes used to categorize an individual as carrying the comorbidity is provided in **Supplementary Table 17**. All odds ratio calculations were conducted using the SciPy v1.11.3 package.

### Mediation analysis

We conducted structural equation modeling (SEM) to explore the complex relationship between gene, BMI, and obesity-associated comorbidities of the BMI-associated genes which are also associated with increased risk of obesity-associated comorbidities. We hypothesized paths from (1) gene → BMI, (2) gene → comorbidity, and (3) gene → BMI → comorbidity. A graphical representation of the path diagram is shown in **Supplementary** Fig. 3. Since our path contains main effects of the gene on the comorbidity and their mediation effect through BMI, we performed a mediation analysis using *lavaan*^50^ to test the most likely path of propagation of disease risk. For each of the four BMI-associated genes which also increased the risk of obesity-associated comorbidities, we fitted SEM model separately in UKB and AoU cohorts to obtain model coefficients and relevant statistics for the specified paths in the path diagram. We then combined the statistics generated from the two cohorts using IVW fixed effect meta-analysis to obtain meta statistics for the coefficients. The meta-analysis was performed using python packages SciPy and NumPy with in-house generated scripts.

### Phenome-wide association study

To identify additional phenotypic associations for the five discovered genes, we conducted a PheWAS across multiple ancestries. ICD-10 diagnosis codes were mapped to phecodes using the PheWAS R package^51^ to create phenotype variables. Gene carrier status was assigned based on the presence of rare PTVs, collapsed by the most deleterious gene burden mask which reached exome-wide significance for that gene. PheWAS was then performed separately for each ancestry using R PheWAS package, adjusting for relevant covariates such as age, age^2^, sex, age × sex and 10 genetic PCs. The analysis was restricted to phenotypes with at least fifty cases, and ancestry-specific associations were combined using IVW fixed effect meta-analysis. Phenotypes that reached a Bonferroni corrected threshold (*P* < 2.7×10^-5^; 1852 phecodes tested) were considered significant associations.

### Interaction model for rare variants and polygenic risk

To assess interactive effects between each gene and PGS, we used a linear regression model to predict BMI using individual as well as interactive terms between PGS and PTV carrier status for the discovered genes while accounting for age, age^2^, sex, age × sex and ten genetic PCs. We obtained separate model coefficients in European individuals of UKB and AoU cohorts and then used IVW fixed effect meta-analysis to combine the statistics from the two cohorts. The models were trained using “ols” function from statsmodels v0.14.2 package, available in python.

### Proteomics data analysis

Normalized plasma protein expression data for 2,923 proteins from 53,058 individuals were obtained through the DNANexus portal of the UKB RAP. For each protein, multiple technical replicates were averaged to generate a single readout per individual. Additional proteomics-related covariates, including the number of proteins tested per individual (Field 30900) and the assay plate used (Field 30901), were extracted for inclusion in the analysis. To assess the impact of PTVs in BMI-associated genes on plasma protein levels, we employed linear regression models, adjusting for age, age^2^, sex, age × sex, age^2^ × sex, number of proteins measured, plate used, genotyping array batch, and 20 genetic PCs. Statistical significance was determined using a Bonferroni-corrected threshold of P < 1.71×10^-5^, accounting for the number of proteins tested (N =2,923). To further evaluate the downstream consequences of significant protein changes, we examined the association between these proteins and normalized BMI using linear regression, adjusting for age, age^2^, sex, age × sex, BMI PGS, exome release batch, and 10 genetic PCs. All models were implemented using the “ols” function from the statsmodels v0.14.2 package in Python.

## AUTHOR CONTRIBUTIONS

S.G and D.B conceived the study, acquired, analyzed and interpreted the data, and wrote the manuscript.

## Data Availability

All data produced in the present work are contained in the manuscript

## ACKNOWLEDGEMENTS

We thank the participants and investigators in the UK Biobank and All of US research studies who made this work possible. This research has been conducted using the UK Biobank Resource under Application Number 45023. We also thank the National Institutes of Health’s All of Us Research Program for making available the participant data examined in this study. This work was supported by NIH R01-GM121907, resources from the Huck Institutes of the Life Sciences and the Pennsylvania State University to S.G. The authors declare no competing interests.

## Data and code availability

The UK Biobank and All of Us genetic and phenotypic data analyzed in this study are publicly available to registered researchers through their respective analysis portals. This study used data from the All of Us Research Program’s Controlled Tier Dataset v8, available to authorized users on the Researcher Workbench. Additional information about registration for access to the data is available at https://www.ukbiobank.ac.uk/ and https://www.researchallofus.org/ for UK Biobank and All of Us, respectively. The code for pre-processing, filtering, annotating genetic data, association tests and statistical analysis is available on GitHub (https://github.com/deeprob/BMI_monogenic).

## Notes

### Competing Interest Statement

The authors have declared no competing interest.

### Funding Statement

This study was funded by the NIH GM121907 grant

### Author Declarations

The data used in this study were obtained after permissions from the UKB and All of Us portals. The Penn State IRB did not consider them as human subjects research.

### Summary of Updates

In this version, we have made the following changes: (1) Expanded our analysis by incorporating 150,000 additional Whole Genome Sequencing (WGS) samples from the latest All of Us release, increasing our total sample size to 839,110. (2) Implemented an ancestry-wise inverse variance-weighted fixed-effect meta-analysis for rare variant gene burden associations. This approach replaces the previously used population-based random-effects meta-analysis, which is more susceptible to population stratification bias and potential false-positive associations. (3) The larger sample size and improved meta-analysis strategy resulted in a substantially smaller number of identified obesity-associated genes compared to our previous findings, suggesting a more refined and robust discovery framework.

