## Supplementary Note for "Discovery of novel obesity genes through cross-ancestry analysis"

### Biobank access for UK Biobank (UKB) and All of Us (AoU)

The UK Biobank and All of Us genetic and phenotypic data analyzed in this study are publicly available to registered researchers through their respective analysis portals. Additional information about registration for access to the data is available at [https://www.ukbiobank.ac.uk/](https://www.ukbiobank.ac.uk/enable-your-research/apply-for-access) and <https://www.researchallofus.org/> for UK Biobank and All of Us, respectively. This research has been conducted using the UK Biobank Resource under Application Number 45023. Data from the All of Us Research Program’s Controlled Tier Dataset v8 was used in this study, available to authorized users on the Researcher Workbench. We thank the participants and investigators in the UK Biobank and All of US research studies who made this work possible. We also thank the National Institutes of Health’s All of Us Research Program for making available the participant data examined in this study.

### Genetic data processing

**Supplementary Table 18**: SNP array, sample and variant quality control by selected publications

| Steps | Backman | Turcot | Akbari | Zhao | Jurgens |
| --- | --- | --- | --- | --- | --- |
| Array QC | - MAF >1% - Missingness <10% - HWE *P>*10^−15^ - LD pruned r^2^ =0.9 | **Not provided**. UKB study specific field is blank in Tables S1-3 | **Not provided**. Methods is sparse, supplementary cites Bycroft UKB paper | - MAF >0.1% - Missingness <10% - HWE *P>*10^-6^ | - MAF >0.1% - Missingness <1% - HWE *P*>10^-6^ |
| Variant QC | - DP>=7 or >=10 (SNV or indel) - At least one homo, or - AB >= 0.15 or 0.2 for SNV or indel - Missingness <10% - HWE *P>*10^−15^ - Not monomorphic | - Missingness <5% - HWE *P>*10^−7^ - Allele frequency deviation from reference populations <0.6 | **Not provided**. | - DP>=7 or >=10 (SNV or indel) - GQ>=20 - Binom test *P>=*0.001 AA vs RA for hetz - Call rate>50% | - 10<=DP<=200 - GQ>=20 - (DP_A1 + DP_A2)/DP > 0.9 - DP_A2/DP > 0.2 or 0.9 for hetz or homo - Call rate > 90% - HWE *P>*10^−15^ - Not in low complexity regions - Not monomorphic |
| Sample QC | - Concordant sex - Low heterozygosity - High coverage - Not duplicates - WES and Array concordance | - Missingness <5% - Referred to UKB pdf – link not working in Table S1. | **Not provided**. | - Low heterozygosity - Array missingness <5% - Phased Bycroft | - Not duplicates - Concordant sex - Missingness <10% - Not deviated from mean Ti/Tv, Het/Hom, SNV/indel, singletons |
| Variant annotation | - Tool: snpEff - pLoF: stop gained, start lost, splice donor, splice acceptor, stop lost, frameshift - Missense: SIFT, PolyPhen2 HDIV and HVAR, LRT, MutationTaster | - Tool: **Not provided** - pLoF: non-sense, stop loss, splice site - Missense: SIFT, PolyPhen2 HDIV and HVAR, LRT, MutationTaster | - Tool: snpEff - pLoF: stop gained, start lost, splice donor, splice acceptor, stop lost, frameshift - Missense: SIFT, PolyPhen2 HDIV and HVAR, LRT, MutationTaster | - Tool: VEP v104 - pLoF: stop gained, splice acceptor, splice donor, frameshift - Missense: REVEL | - Tool: VEP v105 - pLoF: stop gained, splice acceptor, splice donor, frameshift - Missense: 30 tools |
| Burden preparation | - MAF <1, 0.1, 0.01, 0.001% and singletons - “Strict” mask: pLoF - “Permissive” mask: pLoF and del missense by 5/5 tools | - MAF <5% - “Broad” mask: pLoF and del missense by 1/5 tools - “Strict” mask: pLoF and del missense by 5/5 tools | - MAF <1, 0.1% - pLoF only - pLoF or del missense by 5/5 tools - pLoF or del missense by 1/5 tools | - MAF <0.1% - pLoF only - pLoF with missense REVEL>0.7 - pLoF with missense REVEL>0.5 | - MAF <0.1, 0.001% - pLoF only - pLoF and missense score > 0.8 - pLoF and missense score > 0.5 |

#### UKB

SNP array data quality control and processing was performed by following the protocol described by UKB Research team here: ([link](https://dnanexus.gitbook.io/uk-biobank-rap/science-corner/end-to-end-target-discovery-with-gwas-and-phewas/)). In brief, the data is lifted over from GRCh37 to GRCh38 format using a WDL script prepared by Yih-Chii Hwang and shared via Github ([link](https://github.com/dnanexus-rnd/liftover_plink_beds)). The lifted over SNPs went through the following quality control steps: a) Minor allele frequency greater than 0.01, b) Minor allele count greater than 100, c) Missing call rate for variant not exceeding 0.1, d) Missing call rate for sample not exceeding 0.1, e) Hardy-Weinberg equilibrium exact test p-value for the variant greater than 1 x 10^-15^. The filtered SNPs were further LD pruned with an *r*^2^  threshold of 0.9, 1000 variants window size and 100 variant window slide, leaving 480,372 sites that passed all filters. Quality control and LD pruning was performed using PLINK 2.0.

Variant quality control for exome sequencing data entailed filtering both low quality genotype and variant calls. At first, multiallelic sites were split to bi-allelic sites. Genotype calls with genotype quality (GQ) >= 20, depth (DP) >=10 (5 for haploid genotypes on sex chromosomes), and allele balance (AB) >= 0.2 and <= 0.8 (for heterozygous genotypes only) were retained following gnomad guidelines. Subsequently, variants with call rate < 90%, HWE *P*<10^-15^, those present within Ensembl low-complexity regions or monomorphic variants were filtered. Number of variants per chromosome present in the initial pVCF file and after filtering is provided in **Supplementary Table 19**. All computations were performed through Hail in DNA Nexus portal using 40 nodes of “mem2_ssd1_v2_x32” instance type except chromosome 21 where 30 nodes of “mem2_ssd1_v2_x16” were used. In the UK Biobank pVCF files for WES, there were 27,040,643 variants across 469,835 samples and after filtering 19,754,746 were retained.

**Supplementary Table 19**: Per chromosome variant quality control statistics in UK Biobank

| Chromosome | Time taken (hr:min) | Cost (in £) | Initial variants | QC passed variants |
| --- | --- | --- | --- | --- |
| 1 | 3:45 | 35.4 | 2687683 | 1993448 |
| 2 | 2:49 | 26.93 | 1986463 | 1437866 |
| 3 | 2:20 | 22.27 | 1572621 | 1143529 |
| 4 | 1:41 | 16.06 | 1088895 | 758738 |
| 5 | 1:49 | 18.06 | 1200720 | 866217 |
| 6 | 2:04 | 19.73 | 1343340 | 977172 |
| 7 | 2:01 | 19.05 | 1290956 | 934228 |
| 8 | 1:33 | 14.67 | 983421 | 714068 |
| 9 | 1:47 | 16.99 | 1160267 | 855746 |
| 10 | 1:40 | 15.80 | 1105534 | 793288 |
| 11 | 2:22 | 22.53 | 1589236 | 1214865 |
| 12 | 2:13 | 21.19 | 1436008 | 1037880 |
| 13 | 0:57 | 9.08 | 485362 | 344827 |
| 14 | 1:26 | 13.58 | 840039 | 617157 |
| 15 | 1:33 | 14.83 | 936837 | 685335 |
| 16 | 2:00 | 19.09 | 1300373 | 1001211 |
| 17 | 2:21 | 22.42 | 1565173 | 1208075 |
| 18 | 0:52 | 8.35 | 433154 | 309899 |
| 19 | 2:37 | 24.97 | 1791980 | 1348929 |
| 20 | 1:14 | 11.78 | 686927 | 522007 |
| 21 | 1:20 | 5.1 | 289754 | 208899 |
| 22 | 1:06 | 10.63 | 613857 | 467878 |
| X | 1:08 | 10.94 | 652043 | 313484 |

Sample quality control in UKB, we first defined a set of high quality autosomal and X chromosome variants using exome sequencing data. Autosomal variants were filtered for MAF>0.1%, missingness<1%, HWE *P*>10^-6^ and further LD pruned using “indep-pairwise” function with 500 50 0.2 arguments in PLINK to retain a set of high-quality autosomal variants. X-chromosomal variants with missingness < 1%, HWE P>10^-6^, not within pseudo autosomal regions, and LD pruned using “indep-pairwise” function with 500 50 0.2 parameters in PLINK were defined as high-quality X-chromosome variants. These operations were performed in PLINK and not Hail since LD pruning is much faster and economically efficient in PLINK compared to Hail. For example, the autosomal variant filtering step for chromosome 21 in Hail with 40 nodes of “mem2_ssd1_v2_x32” instance type took 42 mins and cost £6.9755 while the same set of operations using a single node of identical instance type in PLINK took 6 mins and cost £0.0604. The number of high-quality variants defined per chromosome is provided in **Supplementary Table 20**. The high-quality LD pruned X chromosome variants were used to infer the genetic sex of each individual using Hail’s “impute sex” function. The autosomal high-quality variants were merged together using plink to create a file with 199,774 high quality variants before LD pruning. We used KING2 algorithm and the high-quality autosomal variants to estimate pairwise kinship coefficients between samples and flagged all third degree or closer relatives based on the coefficients. Since the KING algorithm does not recommend LD pruning, we used the high-quality variants which were not LD pruned to flag related individuals ([link](https://www.kingrelatedness.com/manual.shtml#WITHIN)). We identified 135,287 related and 334,548 unrelated individuals based on KING estimates. Next, we merged the high-quality pruned variants per chromosome into a single file using plink resulting in 129,651 high quality LD pruned autosomal variants. Using the high quality pruned autosomal variants of the unrelated individuals, we first calculated their top 20 genetic principal components (PCs) and then projected the variants of the related individuals into the PC space of the unrelated individuals, using Hail. Additionally, we calculated the heterozygote concordance rates high quality autosomal calls and array calls for all samples. Furthermore, using all autosomal exome variants, we calculated the following sample quality control metrics, a) call rate, b) SNV/indel ratio, c) transition/transversion ratio, d) heterozygote/homozygote ratio, e) insertion/deletion ratio, and f) number of singletons. We then regressed out the 20 ancestral PCs calculated before from quality control metrics b) to f) to calculate ancestry-residualized values for the ratios. Additionally, we extracted the following information about previously flagged samples calculated using array data: a) survey sex (UKB Field: 31), b) genetic sex (UKB Field: 22001), c) sex chromosome aneuploidy (UKB Field: 22019), d) outlier for heterozygosity or missingness (UKB Field: 22027), and e) genetic kinship to other participants (UKB Field: 22021). Finally, we flagged the following samples:

- Missing array data
- Duplicates based on KING2 algorithm
- Sex discordance between survey, exome and array imputations
- Heterozygote concordance rate between high quality autosome and array calls less than 0.8
- Sex chromosome aneuploidy based on array data
- Outliers of heterozygosity or missingness based on array data
- With ten or more third degree relatives
- Less than 90% call rate in exome calls
- More than eight standard deviations from the mean of ancestry residualized SNV/indel ratio, transition/transversion ratio, heterozygote/homozygote ratio, insertion/deletion ratio, and number of singletons.

**Supplementary Table 20**: Per chromosome high quality variant statistics in UK Biobank

| Chromosome | Time taken (hr:min) | Cost (in £) | Initial variants | High QC variants | High QC LD pruned variants |
| --- | --- | --- | --- | --- | --- |
| 1 | 0:49 | 0.49 | 2687650 | 19934 | 12998 |
| 2 | 0:37 | 0.37 | 1890608 | 13906 | 9288 |
| 3 | 0:30 | 0.29 | 1497787 | 11076 | 7121 |
| 4 | 0:22 | 0.22 | 1032712 | 7756 | 5508 |
| 5 | 0:24 | 0.24 | 1200708 | 8752 | 5977 |
| 6 | 0:26 | 0.26 | 1273305 | 10539 | 6455 |
| 7 | 0:28 | 0.21 | 1222490 | 9860 | 6594 |
| 8 | 0:18 | 0.18 | 935307 | 7058 | 4916 |
| 9 | 0:23 | 0.23 | 1100325 | 9091 | 6054 |
| 10 | 0:22 | 0.22 | 1105522 | 8635 | 5815 |
| 11 | 0:30 | 0.3 | 1589220 | 12412 | 7497 |
| 12 | 0:27 | 0.27 | 1363975 | 10217 | 6686 |
| 13 | 0:07 | 0.07 | 460307 | 3476 | 2565 |
| 14 | 0:12 | 0.11 | 840031 | 6330 | 4221 |
| 15 | 0:12 | 0.11 | 936831 | 6547 | 4306 |
| 16 | 0:25 | 0.25 | 1236907 | 10168 | 5990 |
| 17 | 0:28 | 0.28 | 1489257 | 12073 | 7442 |
| 18 | 0:08 | 0.08 | 433149 | 3206 | 2434 |
| 19 | 0:33 | 0.33 | 1791970 | 15367 | 9289 |
| 20 | 0:09 | 0.09 | 686915 | 5644 | 3653 |
| 21 | 0:06 | 0.06 | 289748 | 2610 | 1737 |
| 22 | 0:09 | 0.09 | 613853 | 5117 | 3105 |
| X | 2:15 | 3.06 | 652053 | 604261 | 578596 |

Variant annotation, entailed extracting the quality-controlled variants from the autosomes, filtering for rare variants (MAF<0.01), and annoating the variants using VEP v109 and dbNSFP v4. After annotation, we extracted the gene symbol, transcript id, transcript consequences and biotype of the variant. For variants annotated as missense, we further calculated a deleteriousness score based on nine deleteriousness prediction tools. The score is equivalent to the number of tools that predicted the variant to be deleterious out of the nine tools. After annotations, we only retained variants annotated as stop gained, frameshift, stop lost, start lost, splice acceptor, splice donor and missense variants with a deleteriousness score greater than four. After filtering out the flagged samples during sample quality control, we collected the sample ids of both heterozygous and homozygous samples carrying the retained variants. All computations were performed in the DNANexus portal using Hail, parallelly per chromosome with 30 nodes of instance type “mem2_ssd1_v2_x32” for all except chromosome 1 for which we used 50 nodes of “mem2_ssd1_v2_x32”. The time taken, cost and number of annotated rare variants retained is provided in **Supplementary Table 21**.

**Supplementary Table 21**: Per chromosome annotated rare variant statistics in UK Biobank

| Chromosome | Time taken (hr:min) | Cost (in £) | QC passed variants | Annotated rare PTVs |
| --- | --- | --- | --- | --- |
| 1 | 0:37 | 7.12 | 1993448 | 362638 |
| 2 | 0:36 | 4.29 | 1437866 | 290014 |
| 3 | 0:31 | 3.71 | 1143529 | 245620 |
| 4 | 0:29 | 3.45 | 758738 | 152929 |
| 5 | 0:29 | 3.53* | 866217 | 160597 |
| 6 | 0:30 | 3.60 | 977172 | 181680 |
| 7 | 0:26 | 3.21 | 934228 | 175941 |
| 8 | 0:14 | 1.6* | 714068 | 139558 |
| 9 | 0:27 | 3.19 | 855746 | 148351 |
| 10 | 0:26 | 3.20 | 793288 | 146772 |
| 11 | 0:29 | 3.46 | 1214865 | 237809 |
| 12 | 0:14 | 1.68* | 1037880 | 206029 |
| 13 | 0:23 | 3.92 | 344827 | 66187 |
| 14 | 0:25 | 3.33 | 617157 | 125343 |
| 15 | 0:25 | 2.99 | 685335 | 131745 |
| 16 | 0:27 | 3.27 | 1001211 | 177097 |
| 17 | 0:30 | 3.55 | 1208075 | 229105 |
| 18 | 0:23 | 4.36 | 309899 | 57836 |
| 19 | 0:31 | 3.62 | 1348929 | 213013 |
| 20 | 0:24 | 2.86 | 522007 | 87064 |
| 21 | 0:22 | 2.63 | 208899 | 39261 |
| 22 | 0:23 | 2.75 | 467878 | 82258 |

*These chromosomes shared the overhead cost included in chromosome 5 estimate.

Gene burden preparation was performed by first assigning each variant either of the three labels, a) predicted loss of function (lof), b) missense strict and c) missense lenient, based on their transcript consequence and deleteriousness score annotations. Variants annotated as "frameshift variant", "stop gained", "splice acceptor variant", and "splice donor variant" were assigned the lof label. Missense variants with deleteriousness score of 9 were assigned “missense strict” and those with scores greater than 6 but less than 9 were assigned “missense lenient”. Variants with these three assigned labels and of “biotype” protein coding were retained. Second, we added gnomad exome minor allele frequency annotations of five superpopulations (AFR, AMR, EAS, SAS, EUR) to the retained variants using VEP v109 and Hail. Third, we grouped all variant-gene pairs keeping the highest consequence annotation for that pair (pLoF > missense strict > missense lenient). Additionally, for each variant we assigned an alternate allele frequency based on the maximum of the minor allele frequency in the UKB cohort and the five gnomad annotated superpopulations. Finally, we defined three gene burden grouping masks, a) pLoF: which contains variants annotated as lof for the gene, b) Missense strict: which contains missense variants with deleteriousness score of 9 along with lof variants, and c) Missense lenient: which contains missense variants with deleteriousness score greater than 6 along with missense strict and lof variants.

#### AoU

SNP array data quality control and processing in AoU involved the same steps as followed for UKB. After qc filters and LD pruning, we retained 193,351 variants from array data. The operation was performed in the AoU Researcher workbench using plink v2 with 32 CPUs and 208 GB of RAM and it took ~1 hour to finish.

Variant quality control in AoU team was centrally performed by extensive filtering of low quality genomic data including removal of variants with no high quality genotypes, those that do not pass ExcessHet, QUAL or Allele specific variant quality score recalibration tool scores as detailed here ([link](https://support.researchallofus.org/hc/en-us/articles/29390274413716-All-of-Us-Genomic-Quality-Report)). We used the multiallelic split Hail Matrix table of the exome region provided as smaller srWGS callset by AoU where the low quality sites were already filtered as mentioned here ([link](https://support.researchallofus.org/hc/en-us/articles/14929793660948-Smaller-Callsets-for-Analyzing-Short-Read-WGS-SNP-Indel-Data-with-Hail-MT-VCF-and-PLINK)). In addition to the already performed qc, we further filtered genotypes with quality (GQ)<20 and variants with call rate<90%, HWE *P*<10^-15^, those present within Ensembl low-complexity regions or monomorphic variants. These computations were performed in the AoU Researcher Workbench using Hail and a data processing cluster with 16 CPUs and 104 GB of RAM for the driver and worker nodes. We allocated 4 non-preemptible drivers and 50 preemptible workers for this task, and it took ~40 mins to execute.

Sample quality control in AoU has already been performed by checking and flagging samples that failed quality control measures such as sex discordance, fingerprint discordance, population outliers after regressing out the PCAs from callset metrics such as transition/transversion ratio, or those who are highly related. According to the AoU QC report for data release version 8, no samples with sex discordance were included in the data release. However, some samples who did not respond “male” or “female” in the sex at birth survey question were automatically included. Additionally, all samples included in the release passed fingerprint concordance with array data. Therefore, the flagged samples in AoU were population outliers based on ancestry residualized genetic qc metrics. Apart from filtering out all the flagged samples, we also removed possible duplicates based on AoU provided kinship scores greater than 0.354, and those with sex-at-birth not reported as Male or Female.

Variant annotation in AoU, was performed by following the same steps as described in UKB except the variants were annotated using NIRVANA annotations provided as a Hail table by AoU researcher team and available to use in the AoU Researcher workbench, instead of VEP. Additionally, biotype annotations for transcript consequences of a variant were not provided in the NIRVANA annotation table and hence could not be used.

Gene burden preparation entailed following the same steps as in UKB except variants were not filtered by “protein coding” biotype due to unavailability of the said annotation field. Additionally, gnomad genome minor allele frequency annotations for the five superpopulations available in NIRVANA were used instead of gnomad exome annotations minor allele frequency annotations available through VEP in UKB.

**Note**: The SNP array data quality control and processing protocol described here is ***not*** the same as the one used for ancestry inference. To maintain consistency between UKB and AoU ancestry inference, we followed a different quality control protocol as described in the pipeline for ancestry inference. On the other hand, the SNP array processed through the pipeline described above was used to run Step 1 of the gene burden association analysis in both UKB and AoU.

### Ancestry inference

**Supplementary Table 22**: Ancestry inference steps followed by previous selected studies

| Steps | Backman | Jurgens | AoU |
| --- | --- | --- | --- |
| Reference dataset | HapMap3 | 1K Genome | HGDP and 1K Genome |
| Quality Control | - overlapped SNPs - MAF>10% - Missingness<5% - HWE *P>*10^-5^ | - overlapped SNPs - MAF>1% - Missingness<2% - HWE *P>*10^-4^ | - overlapped SNPs - MAF>0.1% - Missingness<1% - LD pruned r^2^=0.1 |
| PCA computation | Not mentioned | PCAir (N=20) | Hail (N=16) |
| Ancestry prediction | Kernel Density Estimator | ADMIXTURE | Random Forest |

#### UKB

*For ancestry inference of individuals in UKB*, we followed the same protocol as previously used to predict ancestry in AoU. HGDP and 1000 Genome variant data with ancestry labelled for each individuals were used as reference data. HGDP and 1000 Genome samples which passed the “high quality”, “gnomad high quality” and were unrelated based on “relatedness inference related” field were kept resulting in 3,224 individuals. Similarly, UKB samples where sex and genetic sex were the same, there were no sex chromosome aneuploidy, no excessive kinship i.e. not ten or more third degree relatives, not identified as outliers that are heterozygous and missing rates, were kept resulting in 485,932 individuals. We then identified overlapping sites between the UKB genotyped array and the variant data of HGDP and 1000 Genomes samples. These sites were filtered to keep autosomal variants with a minor allele frequency greater than 0.1% and call rate greater than 99% in HGDP, 1000 Genome and UKB datasets. These were further LD pruned with a cut-off of r2=0.1 to obtain a maximal subset of variants that are nearly uncorrelated within a window size of 1000000 bp. The final number of overlapping, high quality sites between the reference and UKB datasets was 238,493. We next generated the first 16 principal components (PCs) of the reference samples using “hwe normalized pca” function in Hail as the feature vector for those samples. Subsequently, we projected the genotypes from the UKB samples into the PC space of the reference samples using “pc project” function in Hail to create the feature vectors for those samples.

To assign ancestries to each individual in UKB, we trained a random forest model using 16 PCs from the reference dataset as the features and their gnomad assigned ancestries as the labels. We used a five-fold cross validation strategy to optimize the max depth and number of classifier hyper parameters. The assigned ancestries by gnomad included: African (afr); South Asian (sas); East Asian (eas); Middle Eastern (mid); Latino/admixed American (amr); European (eur), composed of Finnish (fin) and Non-Finnish European (nfe); Other (oth), not belonging to one of the other ancestries or is an admixture. Individuals assigned to the “oth” label were not used for training. The trained model was used to predict the ancestries of UKB samples using their 16 projected PCs as features. First, we predicted probabilities for the six ancestries of each individual based on their PCs using the random forest model. Individuals with >75% probability for an ancestry were assigned to that ancestry. Individuals with <75% probability for all ancestries were assigned as “oth”.

To assess concordance of the self-reported ethnicity in UKB to the predicted ancestries, we first assigned the “most likely ancestry” to each UKB individuals based on their ethnic background field. We only included those ethnic groups where the ancestry could hypothetically be straightforward to infer namely: “African” to “afr”; “Bangladeshi” to “sas”; “British” to “eur”; “Chinese” to “eas”; “Indian” to “sas”; “Irish” to “eur”; “Pakistani” to “sas”; “White” to “eur”. This resulted in a final group of 442,495 out of 485,932 individuals across four ancestries. Assuming the self-reported ethnicities for individuals correspond to ancestries, the weighted-average precision, recall and f1-score for the four ancestries were 0.999, 0.991 and 0.995 respectively indicating high concordance between the self-reported ethnicity and predicted ancestry (**Supplementary Table 23**). For each self-reported ethnic group, the number of UKB individuals categorized by their predicted ancestry is present in **Supplementary Table 24**.

#### AoU

*Ancestry of individuals was already predicted* and made available publicly by AoU. They used the same protocol as described above for UKB.

**Supplementary Table 23**: Performance of the prediction model using self-reported ethnicities as ground truth

|  | **AFR** | **EAS** | **EUR** | **SAS** | **Accuracy** | **Macro avg** | **Weighted avg** |
| --- | --- | --- | --- | --- | --- | --- | --- |
| **Precision** | 0.999317 | 0.998622 | 0.999986 | 0.998620 | 0.99063 | 0.570935 | 0.999954 |
| **Recall** | 0.931298 | 0.971180 | 0.991485 | 0.970245 | 0.99063 | 0.552030 | 0.990630 |
| **F1-score** | 0.964109 | 0.984709 | 0.995717 | 0.984228 | 0.99063 | 0.561252 | 0.995262 |
| **Support** | 3144 | 1492 | 430398 | 7461 | 0.99063 | 442495 | 442495 |

**Supplementary Table 24**: Ancestry predicted in UKBiobank for each self -eported ethnic groups

| **Ancestry predicted** | **AFR** | **AMR** | **EAS** | **EUR** | **MID** | **OTH** | **SAS** |
| --- | --- | --- | --- | --- | --- | --- | --- |
| **Ethnic background** |  |  |  |  |  |  |  |
| **African** | 2928 | 0 | 0 | 1 | 0 | 214 | 1 |
| **Any other Asian background** | 1 | 0 | 330 | 2 | 7 | 328 | 1041 |
| **Any other Black background** | 88 | 0 | 0 | 0 | 0 | 22 | 4 |
| **Any other mixed background** | 39 | 61 | 4 | 200 | 1 | 614 | 27 |
| **Any other white background** | 0 | 173 | 0 | 12776 | 15 | 2114 | 2 |
| **Asian or Asian British** | 0 | 0 | 2 | 0 | 0 | 14 | 25 |
| **Bangladeshi** | 0 | 0 | 0 | 0 | 0 | 3 | 218 |
| **Black or Black British** | 21 | 0 | 0 | 2 | 0 | 2 | 0 |
| **British** | 1 | 3 | 1 | 414070 | 1 | 3583 | 8 |
| **Caribbean** | 4012 | 1 | 0 | 0 | 0 | 161 | 29 |
| **Chinese** | 0 | 0 | 1449 | 1 | 0 | 41 | 1 |
| **Do not know** | 9 | 8 | 5 | 80 | 2 | 67 | 19 |
| **Indian** | 0 | 0 | 0 | 4 | 0 | 189 | 5331 |
| **Irish** | 0 | 0 | 0 | 12171 | 0 | 37 | 0 |
| **Mixed** | 0 | 2 | 0 | 7 | 0 | 34 | 2 |
| **Other ethnic group** | 756 | 321 | 576 | 438 | 69 | 1587 | 502 |
| **Pakistani** | 0 | 0 | 0 | 0 | 0 | 26 | 1690 |
| **Prefer not to answer** | 144 | 12 | 23 | 1061 | 4 | 158 | 100 |
| **White** | 1 | 2 | 1 | 492 | 0 | 27 | 0 |
| **White and Asian** | 0 | 1 | 3 | 17 | 0 | 695 | 50 |
| **White and Black African** | 27 | 1 | 0 | 6 | 0 | 350 | 1 |
| **White and Black Caribbean** | 55 | 3 | 0 | 6 | 0 | 512 | 2 |
| **Inconsistent** | 139 | 11 | 22 | 12375 | 0 | 369 | 180 |

### Phenotypic data processing

#### UKB

BMI data collection and processing in UKB was performed using Spark JupyterLab instances in the DNANexus portal. Measurements of BMI (Field: 21001) across all assessments for each individual were extracted and mean BMI value across assessments was assigned to each individual.

Covariate data collection and processing in UKB was also performed using Spark JupyterLab instances in the DNANexus portal. The following covariates were extracted:

1. Age (Field: 21003)
2. Sex (Field: 31)
3. Genetic Sex (Field: 22001)
4. Exome release batch (Field: 32050)
5. BMI PGS (Field 26216)
6. Top 10 genetic principal components (Field 22009)
7. Genetic kinship to other participants (Field: 22021)

Age was collected across multiple assessments and mean age was assigned to each individual.

BMI comorbidities collection and processing in UKB involved extracting International Classification of Disease codes (Field: 41270) using Spark JupyterLab instances in the DNANexus portal. For each ICD code, we obtained the samples who were assigned that particular code as well as any lower-level ICD codes which fell under the hierarchy of the original code based on the hierarchical ICD tree. We next obtained the specific ICD codes for obesity associated comorbidities based on previous BMI related studies and annotated individuals as carrying a comorbidity if they were diagnosed with the corresponding ICD codes. A list of the comorbidities and the ICD codes used to categorize an individual as carrying the comorbidity is provided in **Supplementary Table 17.**

Phenotype file preparation in UKB involved aggregating BMI, covariate, comorbidity and lifestyle factor data and further numerically encoding the field as necessary. First, individuals without exome data, those who failed any of the genetic data quality control filters, those without BMI information, those without ancestry predictions or with ancestry predicted as “oth” or those who have ten or more third degree relative were dropped, resulting in a final cohort of 454,645 individuals with exome, phenotype and covariate data. Obesity associated comorbidity information as well as binarized lifestyle factor information was added to the retained individuals as per availability. Others were denoted as “NA” for those fields.

Next, BMI was rank inverse normal transformed [(link)](https://static-content.springer.com/esm/art%3A10.1038%2Fnature11401/MediaObjects/41586_2012_BFnature11401_MOESM14_ESM.pdf), grouped by ancestry and sex of individuals. Number of individuals per ancestry stratified by sex is available in **Supplementary Table 25**. We additionally binarized genetic sex field, assigning value of 1 to females and 0 to males, calculated age^2^ and age x sex terms, flagged individuals who belonged to exome release batch “50K Release” and stored all information in separate files for each ancestry.

**Supplementary Tables 25**: Cohort statistics in UKB stratified by ancestry and sex

|  |  | **Rank inverse normalized BMI** | | | |
| --- | --- | --- | --- | --- | --- |
|  |  | **Mean** | **Min** | **Max** | **N** |
| **Ancestry pred** | **Sex** |  |  |  |  |
| **AFR** | **Female** | 1.7 x 10^-08^ | -3.690671 | 3.690671 | 4471 |
|  | **Male** | 5.47 x 10^-08^ | -3.601525 | 3.601525 | 3161 |
| **AMR** | **Female** | 1.94 x10^-07^ | -3.016449 | 3.016449 | 391 |
|  | **Male** | 9.16 x10^-18^ | -2.797208 | 2.797208 | 194 |
| **EAS** | **Female** | 1.03 x10^-07^ | -3.407804 | 3.407804 | 1527 |
|  | **Male** | -6.26 x10^-08^ | -3.216267 | 3.216267 | 770 |
| **EUR** | **Female** | 3.73 x10^-09^ | -4.599490 | 4.599490 | 236113 |
|  | **Male** | 1.86 x10^-09^ | -4.563970 | 4.563970 | 199222 |
| **MID** | **Female** | 1.79 x10^-17^ | -2.141198 | 2.141198 | 31 |
|  | **Male** | -2.31x 10^-06^ | -2.387809 | 2.387809 | 59 |
| **SAS** | **Female** | -9.59 x10^-08^ | -3.667579 | 3.667579 | 4084 |
|  | **Male** | 1.09 x10^-08^ | -3.699110 | 3.699110 | 4622 |

#### AoU

BMI data collection and processing in AoU was performed in the Researcher Workbench by first creating workspace specific concept sets and datasets which provides a SQL code to extract the selected concepts (Concepts in AoU are analogous to Fields in UKB). We obtained BMI, and BMI measurement date for each sample using the generated SQL code.

Covariate data collection and processing in AoU entailed creating concept sets and datasets with date of birth and sex at birth concepts and extracting them using the dataset generated SQL code in Researcher Workbench. We calculated age of an individual based on the date of birth and the previously extracted BMI measurement date. In addition, 10 genetic PCs and relatedness samples flagged individuals were obtained from the auxiliary genomic quality control files made available by AoU.

BMI comorbidities collection and processing in AoU involved extracting samples who have positive diagnosis for relevant ICD codes for obesity associated comorbidities (**Supplementary Table 17**).

Phenotype file preparation in AoU involved aggregating BMI, covariate, and comorbidity data and further numerically encoding the field as necessary. First, individuals without whole genome data, those who failed any of the genetic data quality control filters, without BMI information, without ancestry predictions or with ancestry predicted as “oth”, those who are related based on AoU quality control data, those with sex at birth not listed as either Male or Female, and samples with BMI values outside the range of 12 to 75 kg/m^2^ as observed in UKB cohort were removed, resulting in a final cohort of 384,465 individuals with exome, phenotype and covariate data.

Next, BMI was rank inverse normal transformed [(link)](https://static-content.springer.com/esm/art%3A10.1038%2Fnature11401/MediaObjects/41586_2012_BFnature11401_MOESM14_ESM.pdf), grouped by ancestry and sex of individuals. Number of individuals per ancestry stratified by sex is available in **Supplementary Table 26**. We additionally binarized genetic sex field, assigning value of 1 to females and 0 to males, calculated age^2^ and age x sex terms, and stored all information in separate files for each ancestry.

**Supplementary Tables 26**: Cohort statistics in AoU stratified by ancestry and sex

|  |  | **Rank inverse normalized BMI** | | | |
| --- | --- | --- | --- | --- | --- |
|  |  | **Mean** | **Min** | **Max** | **N** |
| **Ancestry pred** | **Sex** |  |  |  |  |
| **AFR** | **Female** | 3 x 10^-6^ | -4.091100 | 4.091100 | 46584 |
|  | **Male** | 8 x 10^-6^ | -4.167304 | 4.167304 | 32444 |
| **AMR** | **Female** | 4 x 10^-6^ | -4.253226 | 4.253226 | 47458 |
|  | **Male** | 5 x 10^-6^ | -4.101892 | 4.101892 | 24403 |
| **EAS** | **Female** | 1,7 x 10^-5^ | -3.761721 | 3.761721 | 5926 |
|  | **Male** | 8 x 10^-6^ | -3.625249 | 3.625249 | 3464 |
| **EUR** | **Female** | 5 x 10^-6^ | -4.474012 | 4.474012 | 130267 |
|  | **Male** | 4 x 10^-6^ | -4.387750 | 4.387750 | 87314 |
| **MID** | **Female** | 1.25 x 10^-4^ | -3.011757 | 3.216267 | 770 |
|  | **Male** | 8 x 10^-6^ | -3.173957 | 3.173957 | 665 |
| **SAS** | **Female** | 2 x 10^-5^ | -3.564872 | 3.564872 | 2747 |
|  | **Male** | 8 x 10^-6^ | -3.531819 | 3.531819 | 2423 |

### Gene burden association test across ancestries and biobanks

#### UKB

*Gene burden association tests* in UKB was conducted independently for each ancestry using REGENIE v3.5. *In step 1*, REGENIE fits a null model that estimates a polygenic risk for the given traits using genome-wide SNP data. We used the quality-controlled SNP array file to create the null model with rank inverse normal transformed BMI and untransformed BMI as the traits accounting for age, age^2^, sex, age x sex, exome release batch and first ten genetic PCs as covariates. The computations were performed using the Swiss-army knife tool available in DNANexus portal with “mem1_ssd1_v2_x36” instance type and took 1 hour 30 mins to execute for each ancestry.

In step 2, REGENIE calculates associations between the genetic variants of interest and the trait after considering the null model calculated in step 1 and other user defined covariates. REGENIE is capable of collapsing variants to a gene-level with user defined masks or collapsing models as input files and then calculate the association of the gene mask instead of individual variants. We supplied our already annotated variant file based on their impact on gene transcripts and defined three variant masks to collapse variants on a gene level, namely, (a) “pLoF” variants only, (b) “pLoF” and “Missense strict” variants, and (c) “pLoF”, “Missense strict” and “Missense lenient” variants. Additionally, the AAF of the variants to be considered within a gene mask could also be specified. We provided the previously assigned AAF of the variants calculated based on the maximum of cohort frequency and gnomad annotated superpopulation frequencies as input. We performed independent gene-based rare variant association tests for all six continental ancestries and generated statistics for all individual gene-mask pairs considering variants with a maximum AAF of 0.001. The computations were parallelized by chromosome and performed using the Swiss-army knife tool available in DNANexus portal with “mem1_ssd1_v2_x4” instance type and took approximately 6 mins to execute for each chromosome.

#### AoU

*Gene burden association tests* in AoU was conducted independently for each ancestry using REGENIE v3.3. The same steps were followed in AoU as in UKB except the release batch was not added as a covariate. The computations were performed using JupyterLab session in the AoU researcher workbench. The time estimates were very similar to UKB.

### Meta analysis of independent gene burden association tests

In this study, we performed 12 independent gene burden association test across six ancestries (AFR, AMR, EAS, EUR, MID, and SAS) and two biobanks (UKB and AoU). We combined the statistics into three meta statistics using inverse variance weighted fixed effects model. Therefore, for each gene-mask pair, we obtained three meta statistics:

1. **European meta statistic**: Obtained by combining results from AoU European and UKB European independent association studies.
2. **Non-European meta statistic**: Obtained by combining results from AoU African, AoU American, AoU East Asian, AoU Middle Eastern, AoU South Asian, UKB African, UKB American, UKB East Asian, UKB Middle Eastern, and UKB South Asian independent association studies.
3. **All-ancestry meta statistic**: Obtained by combining results from AoU African, AoU American, AoU East Asian, AoU European, AoU Middle Eastern, AoU South Asian, UKB African, UKB American, UKB East Asian, UKB European, UKB Middle Eastern, and UKB South Asian independent association studies.

Our Bonferroni significance threshold (*P* < 8.33×10^-7^) was defined for 20,000 genes and three variant collapsing masks (“pLoF”, “Missense strict” and “Missense lenient”) tested per gene. Any gene-mask pair which crossed the Bonferroni threshold in “European meta statistic” **and** in “All ancestry meta statistics” was considered to be significantly associated with BMI.
